# Meta-analysis of over 8,000 individuals from Hawai‘i and Samoa for genetic associations to cardiometabolic phenotypes

**DOI:** 10.64898/2026.05.08.26352761

**Authors:** Bryan L. Dinh, Xinran Wang, Xin Sheng, Peggy Wan, Amit Kumar Srivastava, Take Naseri, Satupa‘itea Viali, Lynne Wilkens, Loïc Le Marchand, Christopher Haiman, Daniel E. Weeks, Charleston W.K. Chiang, Jenna C. Carlson

## Abstract

Although genome-wide association studies (GWAS) now routinely reveal genetic associations and biological insights in millions of individuals, underrepresentation of global populations, such as those from Polynesia, continue to persist. These exclusions, often driven by logistical challenges and lack of data, prevent systematic identification of population-enriched associations, such as the association of the missense variant at the CREBRF locus to BMI and type 2 diabetes discovered commonly occurring in Polynesian populations due to its rarity in global populations. Armed with the recently updated TOPMed imputation panel that could benefit studies in diverse populations that previously had poorer imputation performance, we performed the first GWAS of Native Hawaiians and largest to date of Polynesian-ancestry populations (combined N up to 8,461) to identify population-enriched associations for 13 adiposity and cardiometabolic traits available across both cohorts: BMI, fasting glucose, fasting insulin, HDL, height, hip circumference, HOMA-IR, LDL, T2D, total cholesterol, triglycerides, waist circumference, and waist-hip ratio. We found 25 trait-loci associations that met genome-wide significance: 20 previously reported or known associations and 5 associations newly confirmed via meta-analysis. In particular, with improved statistical power, we were able to confirm the suspected association between the missense *CREBRF* variant with fasting glucose levels. The remaining 4 potentially novel loci-trait associations for BMI, LDL, and waist-hip ratio, however, were not replicated in multi-ethnic datasets from All-of-Us despite having reasonable power to replicate. The lack of Polynesian-enriched findings outside of the *CREBRF* locus informs the bounds of the effect sizes or frequency of any enriched variants, and suggests that further expansion of cohort sizes from this region of the world and improved imputation references specific to these populations are needed to identify more population-enriched associations.

## Introduction

Native Hawaiians and Samoans are underserved populations with Polynesian ancestry that experience different incidence rates of BMI, type 2 diabetes (T2D), cancers, and cardiovascular disease (CVD) among others, compared to other continental populations (1–9). As data are scarce and available cohorts are small, they are often excluded from investigations of the genetic or environmental underpinnings of complex traits and diseases. Additionally, the lack of adequate genomic resources (both in available microarray data and genotype imputation references) and complex population structure and admixture history have also complicated their inclusion in genetic investigations (7,10–12). This presents a particular challenge for identifying associations driven by population-enriched alleles – alleles that are tens if not hundreds of folds more common in Polynesia compared to the rest of the world and contribute disproportionately to phenotypic variation and disease risk in Hawaiians, Samoans, and other Pacific Islanders (13–16). As these variants are rare outside of Polynesia, they can only be discovered efficiently with cohorts derived from this region.

While both Native Hawaiians and Samoans represent Polynesian ancestries at the macro-geographical level, there are fine-scale differences between the two populations. Specifically, Native Hawaiians represent eastern Polynesian-related ancestries and, due to European colonization and admixture, at minimum additionally incorporate East Asian-, European-, and African-related ancestries (17). By comparison, Samoans are less admixed and represent predominantly western Polynesian-related ancestries (18). Nevertheless, the two populations as well as other Pacific Islander populations have a shared history and genetic architecture. For instance, a Polynesian-enriched missense allele of the variant rs373863828 (chr5:173108771) in *CREBRF* was first discovered in a Samoan cohort and was common among Native Hawaiian and other Pacific Islander populations (∼9-29%) with similar effect sizes on BMI (14,15,19–22). This allele is virtually non-existent in other continental ancestries (frequency < 2×10^−4^ in gnomAD (23)). Some studies have also suggested that this *CREBRF* missense variant may be associated with other adiposity or metabolic phenotypes, though the statistical evidence is marginal due to lack of power (14,15). Further, a multiple cohort study including Samoans and other individuals of Pacific Islander ancestry discovered another Polynesian-specific missense *CETP* variant, rs1597000001 (chr16:56971035), associated with High Density Lipoprotein Cholesterol (HDL-C) and Low Density Lipoprotein Cholesterol (LDL) levels (16). Hence, combining both populations should improve statistical power to discover additional population-enriched associations shared across Polynesia.

More importantly, since the reporting of those studies, publicly available imputation panels such as the state-of-the-art TOPMed imputation server have improved genotype imputation accuracy through increases in the number of variants (>445 million), number of individuals (>133k), and diversity of representation (24,25), with benefits already observed for Latino and African American populations. In particular, the previously unimputable *CREBRF* variant (15) can now be imputed using TOPMed references to high accuracy, bringing optimism that other Polynesian-enriched variants may now also be better imputed with this reference panel.

In our current study, we imputed both the Native Hawaiians (max N = 5,376) from the Multiethnic Cohort (MEC) study and Samoans (max N = 3,085) from the Soifua Manuia study using release 3 of the TOPMed imputation reference panel and performed genome-wide meta-analysis for several cardiometabolic traits. Overall, these improvements, coupled with the increased sample size from meta-analyzing Native Hawaiian and Samoan cohorts, allow us to confirm previous suggested associations and also improves power to detect other Polynesian-enriched variants.

## Results

Native Hawaiian and Samoan cohorts were meta-analyzed following association testing. Briefly, genotype array data for each cohort were phased and imputed using the TOPMed imputation server (release 3). After quality control filtering, linear mixed model-based association testing for each of the 13 phenotypes (**Table 1, Supplementary Fig 1-2**) was performed via GENESIS followed by meta-analysis via METAL. In total, we identified 775 genome-wide significant variants at 25 loci across the autosomes and chromosome X (**Fig 1, Supplementary Fig 3**).

**Fig 1.**
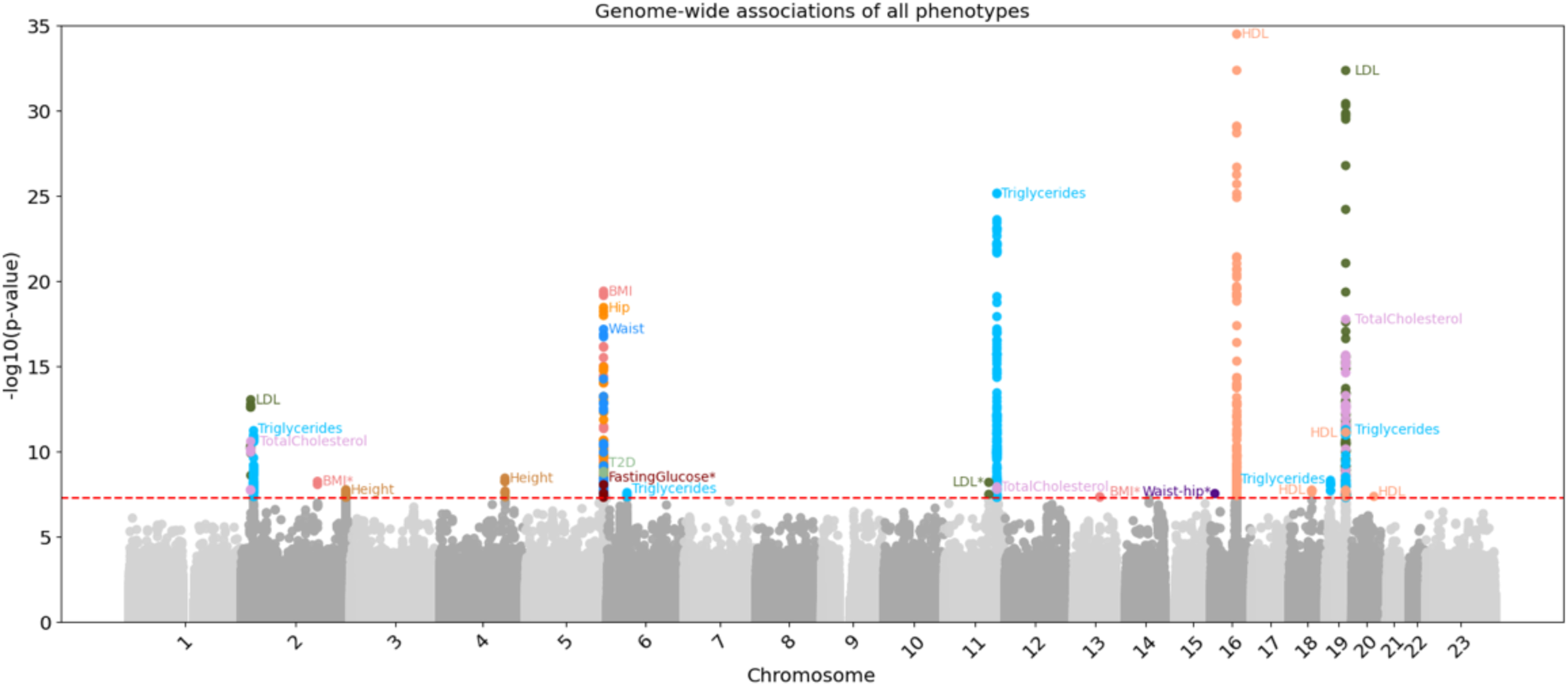
Composite Manhattan plot of meta-analysis associations across all 13 traits. Associations meeting a cutoff of 5e-8 included loci for the following phenotypes: BMI, fasting glucose, HDL, height, hip circumference, LDL, T2D, total cholesterol, triglycerides, waist circumference, and waist-hip ratio. No loci were found to be significant for fasting insulin or HOMA-IR phenotypes.

**Table 1.** Sample availability for traits analyzed in this study. BMI, body mass index; HDL high-density lipoprotein; HOMA-IR, homeostatic model assessment for insulin resistance; LDL, low-density lipoprotein; T2D, type 2 diabetes.

| Phenotype | N |  |  |
| --- | --- | --- | --- |
|  | NH | Samoans | Total |
| **BMI** | 5,368 | 3,083 | 8,451 |
| **Fasting Glucose** | 1,756 | 2,397 | 4,153 |
| **Fasting Insulin** | 1,762 | 2,396 | 4,158 |
| **HDL** | 1,900 | 2,858 | 4,758 |
| **Height** | 5,376 | 3,085 | 8,461 |
| **Hip Circumference** | 3,965 | 3,075 | 7,040 |
| **HOMA-IR** | 1,725 | 2,396 | 4,121 |
| **LDL** | 1,888 | 2,824 | 4,712 |
| **T2D** | 5,343 | 2,875 | 8,218 |
| **Total Cholesterol** | 1,915 | 2,859 | 4,774 |
| **Triglycerides** | 1,896 | 2,825 | 4,721 |
| **Waist Circumference** | 3,999 | 3,074 | 7,073 |
| **Waist-hip ratio** | 3,939 | 3,069 | 7,008 |

We classified each phenotype-locus pair as either previously confirmed (replicating a genome-wide significant finding reported in literature or GWAS catalog (26)) or newly confirmed (not previously reported reaching a genome-wide significance threshold of 5e-8). The 25 loci-trait associations were classified as 5 newly confirmed locus-trait pairs (**Fig 2**, **Table 2**) and 20 previously confirmed locus-trait pairs (**Fig 3**, **Supplementary Fig 4**, **Table 3**). When shown, the recombination rates at each locus reflect inferred rates previously reported for Native Hawaiians in hg38 (27). We named each of the 25 loci by the name of the putative causal gene in each locus, suggested through an artificial intelligence-based literature search (**Methods**; **Tables 2** and **3**). Among the 5 newly confirmed locus-trait pairs, 1 is located at the *CREBRF* locus and associated with fasting glucose levels. The remaining 4 newly confirmed pairs consist of associations for waist-hip ratio (*RBFOX1*, including both sexes), LDL (*CNTN5*), and BMI (*MYCBP2* and *BBS5*). For each locus-trait pair we also performed conditional analysis to discover secondary signals at these loci but found no additional variants reaching genome-wide significance (**Supplementary Table 1**). For the association with fasting glucose levels at the *CREBRF* locus, we performed a secondary conditional analysis by including the known nearby missense variant (rs373863828, chr5:173108771) and verified that our top associations reflect the same signal and that there are no additional signals at this locus (**Supplementary Table 1**).

**Fig 2.**
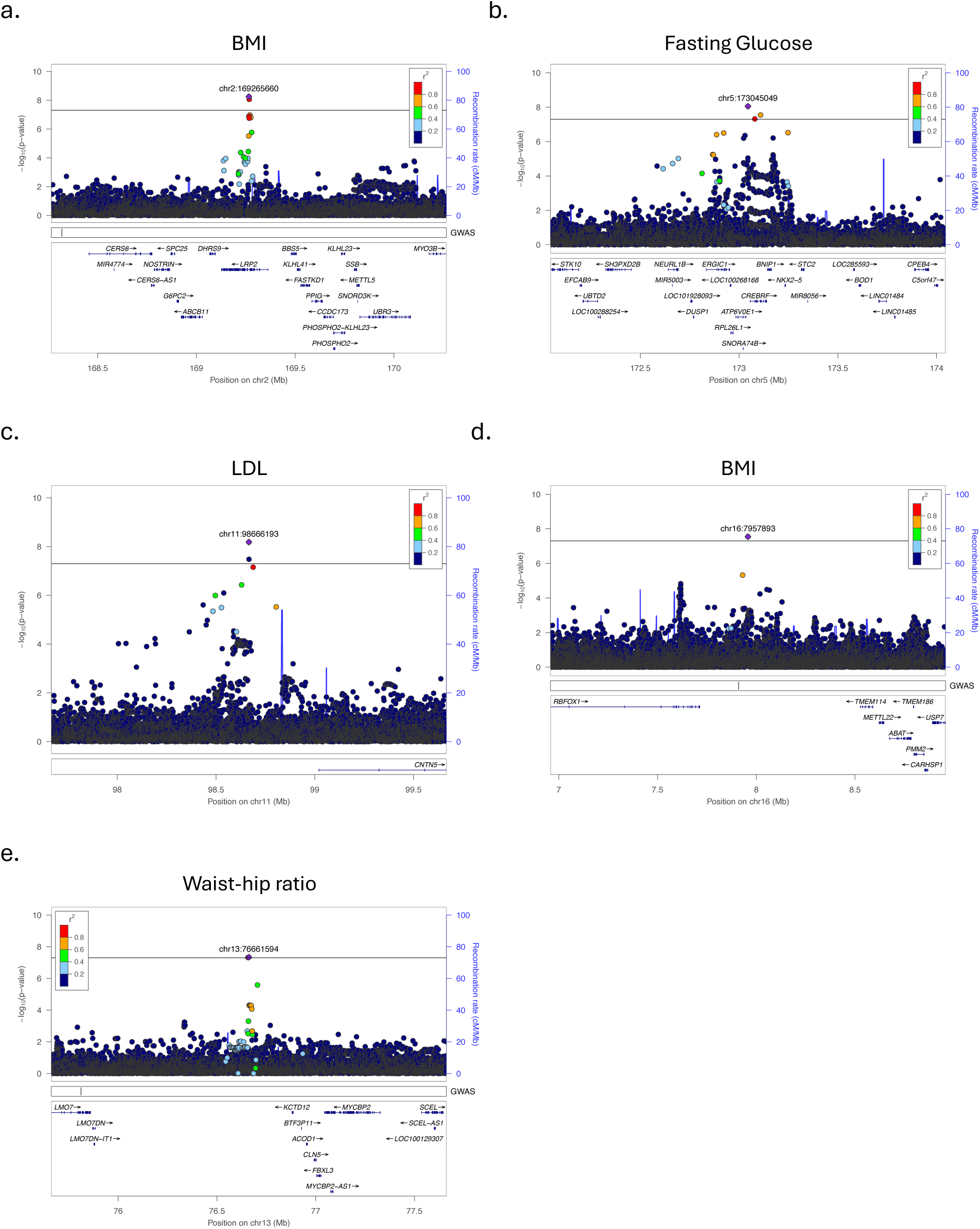
LocusZoom plots for loci categorized as newly confirmed. a) *BBS5* locus for BMI (chr2), b) *CREBRF* locus for fasting glucose (chr5), c) *CNTN5* locus for LDL (chr11), d) *MYBCP2* locus for BMI (chr13), e) *RBFOX1* locus for waist-hip ratio (chr16). The GWAS track, if present, shows reported associations in GWAS catalog for the same trait. Lead variants outside the *CREBRF* locus (a-d) are rarer (allele frequencies ∼0.0017-0.0228) and imputed slightly poorer (mean Rsq 0.89 and 0.78 for Native Hawaiians and Samoans, respectively) compared to the known *CREBRF* missense variant (Rsq 0.991-0.994, AF 0.086-0.29). Further investigation is required to determine if these loci are technical artifacts or possibly tagging rare, unobserved variants in our cohorts.

**Fig 3.**
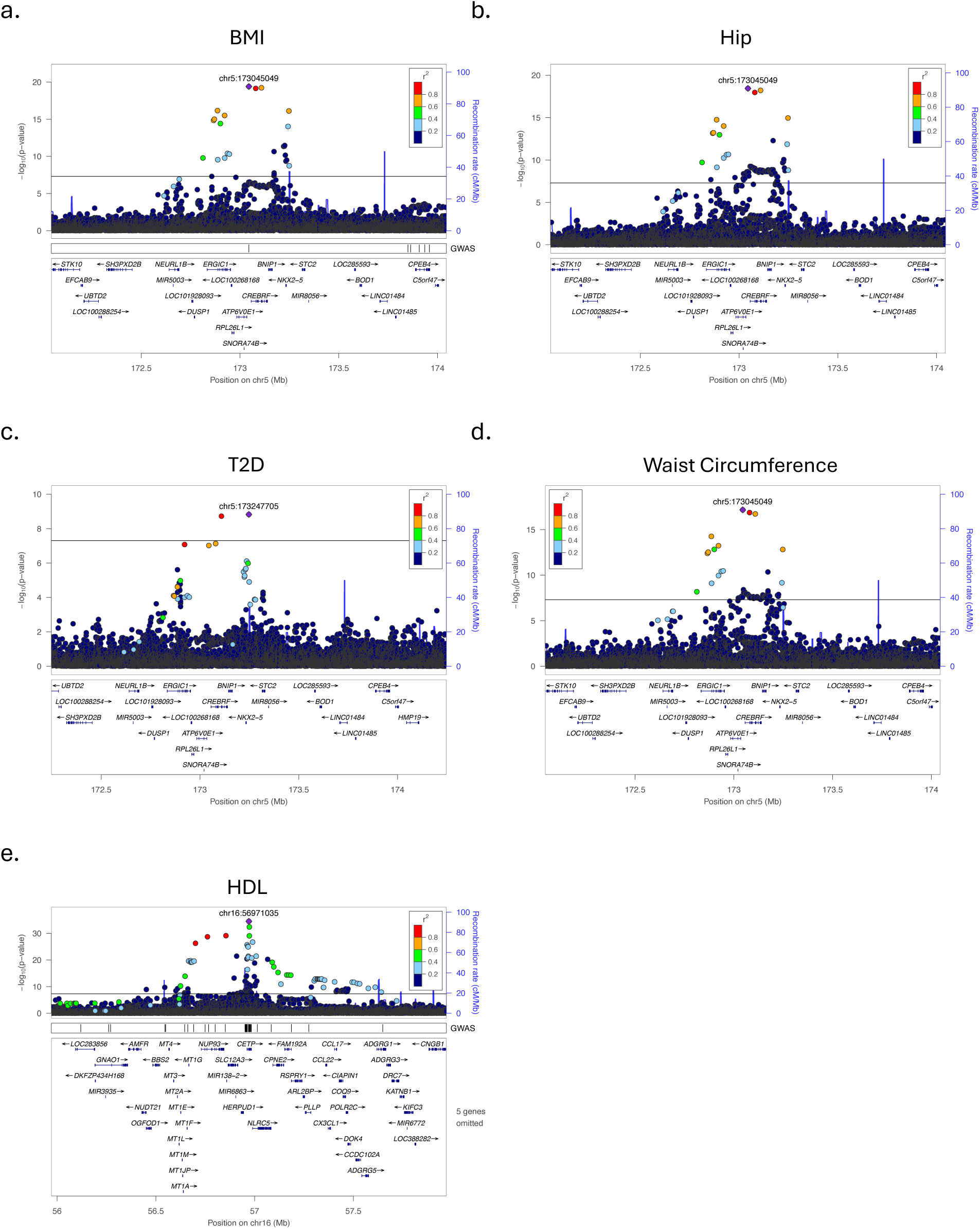
LocusZoom plots for selected loci categorized as previously confirmed. a) *CREBRF* locus for BMI (chr5), b) *CREBRF* locus for hip circumference (chr5), c) *CREBRF* locus for T2D (chr5), d) *CREBRF* locus for waist circumference (chr5), e) *<u>CETP</u>* locus for HDL (chr16). Remaining replicated loci may be found in **Supplementary** Fig 4. The GWAS track, if present, shows reported associations in GWAS catalog for the same trait. Both genes are known to be Polynesian-enriched. The *CREBRF* missense variant is pleiotropic and its impact on BMI, waist (abdominal) circumference, hip circumference, and T2D have previously been reported. A Polynesian-enriched *CETP* variant has previously been associated with HDL.

**Table 2.**
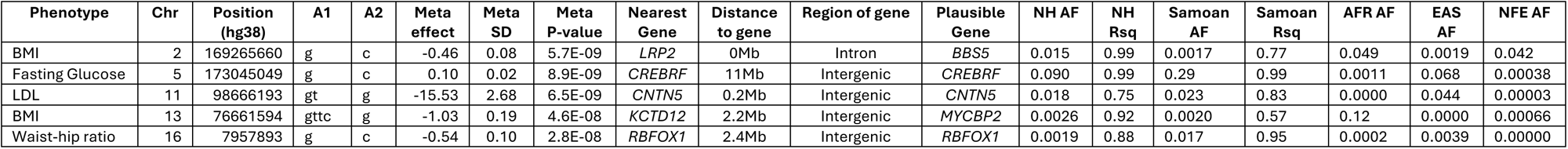
Summary associations for the loci that are identified as newly confirmed. Lead variant in *CREBRF* was found to be newly genome-wide significantly associated with fasting glucose via meta-analysis. Four additional associations for BMI, LDL, and waist-hip ratio also surpassed genome-wide significance but require additional investigation. AI-assisted literature search was used to suggest a plausible associated gene at each locus (**Methods**). NH, Native Hawaiians; AFR, African genetic ancestry group; EAS, East Asian genetic ancestry group; NFE, non-Finnish European genetic ancestry group; AF, allele frequency of the A1 allele; Rsq, imputation r-squared metric for imputation quality. AFR, EAS, and NFE frequency information were retrieved from gnomAD.

| Phenotype | Chr | Position (hg38) | A1 | A2 | Meta effect | Meta SD | Meta P-value | Nearest Gene | Distance to gene | Region of gene | Plausible Gene | NH AF | NH Rsq | Samoan AF | Samoan Rsq | AFR AF | EAS AF | NFE AF |
| --- | --- | --- | --- | --- | --- | --- | --- | --- | --- | --- | --- | --- | --- | --- | --- | --- | --- | --- |
| BMI | 2 | 169265660 | g | c | -0.46 | 0.08 | 5.7E-09 | *LRP2* | 0Mb | Intron | *BBS5* | 0.015 | 0.99 | 0.0017 | 0.77 | 0.049 | 0.0019 | 0.042 |
| Fasting Glucose | 5 | 173045049 | g | c | 0.10 | 0.02 | 8.9E-09 | *CREBRF* | 11Mb | Intergenic | *CREBRF* | 0.090 | 0.99 | 0.29 | 0.99 | 0.0011 | 0.068 | 0.00038 |
| LDL | 11 | 98666193 | gt | g | -15.53 | 2.68 | 6.5E-09 | *CNTN5* | 0.2Mb | Intergenic | *CNTN5* | 0.018 | 0.75 | 0.023 | 0.83 | 0.0000 | 0.044 | 0.00003 |
| BMI | 13 | 76661594 | gttc | g | -1.03 | 0.19 | 4.6E-08 | *KCTD12* | 2.2Mb | Intergenic | *MYCBP2* | 0.0026 | 0.92 | 0.0020 | 0.57 | 0.12 | 0.0000 | 0.00066 |
| Waist-hip ratio | 16 | 7957893 | g | c | -0.54 | 0.10 | 2.8E-08 | *RBFOX1* | 2.4Mb | Intergenic | *RBFOX1* | 0.0019 | 0.88 | 0.017 | 0.95 | 0.0002 | 0.0039 | 0.00000 |

**Table 3.** Summary associations for the loci that replicate previously reported associations. Among the associations, we replicate known Polynesian-enriched associations with BMI (*CREBRF*) and HDL (*CETP*). AI-assisted literature search was used to suggest a plausible associated gene at each locus (**Methods**). NH, Native Hawaiians; AFR, African genetic ancestry group; EAS, East Asian genetic ancestry group; NFE, non-Finnish European genetic ancestry group; AF, allele frequency of the A1 allele; Rsq, imputation r-squared metric for imputation quality. AFR, EAS, and NFE frequency information were retrieved from gnomAD.

| Phenotype | rsid | Chr | Position (hg38) | A1 | A2 | Meta effect | Meta SD | Meta P-value | Nearest Gene | Distance to gene | Region of gene | Plausible Gene | NH AF | NH Rsq | Samoan AF | Samoan Rsq | AFR AF | EAS AF | NFE AF |
| --- | --- | --- | --- | --- | --- | --- | --- | --- | --- | --- | --- | --- | --- | --- | --- | --- | --- | --- | --- |
| Total Cholesterol | rs187411610 | 2 | 21167346 | a | g | 22.5 | 3.381 | 2.6E-11 | *TDRD15* | 0.02Mb | Intergenic | *APOB* | 0.0035 | 0.97 | 0.019 | 0.98 | 0.0000 | 0.0004 | 0.0000 |
| LDL | rs187411610 | 2 | 21167346 | a | g | 23 | 3.085 | 9.3E-14 | *TDRD15* | 0.02Mb | Intergenic | *APOB* | 0.0036 | 0.97 | 0.019 | 0.98 | 0.0000 | 0.0004 | 0.0000 |
| Triglycerides | rs780093 | 2 | 27519736 | t | c | 0.07 | 0.011 | 6.0E-12 | *GCKR* | 0Mb | Intron | *GCKR* | 0.63 | 1.00 | 0.66 | 0.94 | 0.86 | 0.51 | 0.60 |
| Height | rs7571816 | 2 | 232212354 | a | g | 0.09 | 0.016 | 1.8E-08 | *DIS3L2* | 0Mb | Intron | *PRSS56* | 0.30 | 1.00 | 0.37 | 1.00 | 0.098 | 0.50 | 0.022 |
| Height | rs541082621 | 4 | 144412450 | a | g | -0.261 | 0.044 | 3.7E-09 | *LOC105377462* | 0Mb | Intron | *HHIP* | 0.0057 | 0.97 | 0.080 | 0.97 | 0.0000 | 0.0023 | 0.0000 |
| BMI | rs12513649 | 5 | 173045049 | c | g | -0.21 | 0.023 | 3.9E-20 | *CREBRF* | 11Mb | Intergenic | *CREBRF* | 0.088 | 0.99 | 0.28 | 0.99 | 0.0011 | 0.068 | 0.0004 |
| Hip circumference | rs12513649 | 5 | 173045049 | c | g | -0.22 | 0.02 | 3.6E-19 | *CREBRF* | 11Mb | Intergenic | *CREBRF* | 0.086 | 0.99 | 0.28 | 0.99 | 0.0011 | 0.068 | 0.00038 |
| Waist circumference | rs12513649 | 5 | 173045049 | c | g | -0.21 | 0.02 | 6.8E-18 | *CREBRF* | 11Mb | Intergenic | *CREBRF* | 0.086 | 0.99 | 0.28 | 0.99 | 0.0011 | 0.068 | 0.00038 |
| T2D | rs375401688 | 5 | 173247705 | a | g | 0.39 | 0.064 | 1.5E-09 | *CREBRF* | 11Mb | Intergenic | *CREBRF* | 0.061 | 0.95 | 0.25 | 0.89 | 0.0000 | 0.0006 | 0.0000 |
| Triglycerides | rs9472143 | 6 | 43858616 | t | c | -0.067 | 0.012 | 2.6E-08 | *LOC105375070* | 0Mb | Intron | *VEGFA* | 0.23 | 0.99 | 0.27 | 0.88 | 0.14 | 0.12 | 0.24 |
| Total Cholesterol | rs3741298 | 11 | 116786845 | t | c | -4.593 | 0.806 | 1.2E-08 | *ZPR1* | 0Mb | Intron | *APOA5* | 0.51 | 1.00 | 0.37 | 1.00 | 0.66 | 0.59 | 0.80 |
| Triglycerides | rs662799 | 11 | 116792991 | a | g | -0.114 | 0.011 | 7.1E-26 | *APOA5* | 0Mb | 2KB Upstream | *APOA5* | 0.76 | 1.00 | 0.60 | 0.98 | 0.88 | 0.70 | 0.93 |
| HDL | rs1597000001 | 16 | 56971035 | t | c | 8.97 | 0.725 | 3.3E-35 | *CETP* | 0Mb | Exon | *CETP* | 0.017 | 1.00 | 0.035 | 0.96 | - | - | - |
| HDL | rs117998095 | 18 | 50030280 | a | g | -5.486 | 0.976 | 1.9E-08 | *MYO5B* | 0Mb | Intron | *LIPG* | 0.010 | 0.96 | 0.020 | 0.98 | 0.0002 | 0.036 | 0.0000 |
| Triglycerides | rs762055973 | 19 | 11284678 | t | tc | -0.163 | 0.028 | 5.0E-09 | *TSPAN16* | 12Mb | Intergenic | *ANGPTL8* | 0.027 | 0.99 | 0.040 | 0.91 | 0.0000 | 0.0008 | 0.0000 |
| HDL | rs429358 | 19 | 44908684 | t | c | 2.16 | 0.316 | 7.5E-12 | *APOE* | 0Mb | Exon | *APOE* | 0.25 | 0.99 | 0.22 | 0.87 | 0.22 | 0.098 | 0.14 |
| Triglycerides | rs157595 | 19 | 44922203 | a | g | -0.078 | 0.011 | 5.3E-12 | *APOC1* | 0.02Mb | Intergenic | *APOC4* | 0.59 | 0.95 | 0.48 | 0.71 | 0.85 | 0.43 | 0.61 |
| LDL | rs141622900 | 19 | 44923535 | a | g | -19.7 | 1.645 | 4.4E-33 | *APOE* | 0Mb | Exon | *APOE* | 0.057 | 1.00 | 0.065 | 0.66 | 0.081 | 0.084 | 0.056 |
| Total Cholesterol | rs141622900 | 19 | 44923535 | a | g | -15.71 | 1.791 | 1.8E-18 | *APOE* | 0Mb | Exon | *APOE* | 0.058 | 1.00 | 0.066 | 0.66 | 0.081 | 0.084 | 0.056 |
| HDL | rs188671999 | 20 | 48202897 | t | c | 21.5 | 3.923 | 4.3E-08 | *LOC105372641* | 0Mb | 500B Downstream | *NCOA3* | 0.0012 | 0.95 | 0.0012 | 0.83 | 0.0003 | 0.0000 | 0.0032 |

Among previously known loci-trait pairs, five loci were associated with triglycerides (*ANGPTL8*, *VEGFA*, *APOC4*, *GCKR*, *APOA5*), four with HDL (*NCOA3*, *LIPG*, *CETP*, *APOE*), three with total cholesterol (*APOB*, *APOE*, *APOA5*), two with height (*HHIP*, *PRSS56*) and with LDL (*APOB*, *APOE*), and one with BMI, hip circumference, waist circumference, and T2D (all 4 with *CREBRF*). Each of these loci had at least one reported association within 500kb of the top variant reported in the GWAS catalog or in literature for the same phenotype. In each case, the observed top association in our study would be strongly attenuated (12 of 20 loci-trait pairs are no longer nominally significant, *P* > 0.05) if we tested an association model including the previously known variant from GWAS catalog (**Supplemental Table 2**), thereby confirming our observed association reflects the previously reported associations. The five previously known Polynesian-enriched associations at *CREBRF* (for BMI, waist circumference, hip circumference, T2D) and *CETP* (for HDL) were also identified in the meta-analysis, with no other lead variants at these known loci showing obvious enrichment among Polynesian populations (**Table 3**). We note that *CETP* locus was previously shown to be associated with HDL and LDL, but in our meta-analysis the association for LDL was marginal (*P* = 0.007238).

As no other genetic data from Polynesians are available at scale, we sought to replicate our potentially novel associations using the multi-ethnic samples in the All-of-Us Research Program (AoU) (28) when the same or related phenotypes were available in a subset of the AoU subpopulations. We attempted *in-silico* replication of the lead variants in our meta-analysis, or variants in linkage disequilibrium (LD) in our data, across AoU subpopulations as data are available. In general, we observed no replication at nominal significance (at significance level of *P* = 0.01) in any of the variants available (**Supplementary Table 3**). For the associations at *RBFOX1*, *MYCBP2*, and *BBS5*, they are relatively common in other continental ancestries (maximum AF in gnomAD AFR, EAS, and NFE 0.0039-0.12). For these loci, we observed no significant association in AoU to waist circumference, BMI (*MYCBP2*), and BMI (*BBS5*), respectively, despite having substantial statistical power to replicate (max Power = 90.07%, 100% and 100% in AoU SAS, AFR/AMR/EUR, and AFR/AMR/EUR/SAS, respectively; **Supplementary Table 3**). Combined with the fact that these variants appear to be quite rare in Polynesians (0.0019-0.0151 in NH, 0.0017-0.0168 in Samoans), we concluded that they are likely technical artifacts, possibly due to imputation errors, numerical errors in the linear mixed model, or very rare variants that happened to occur in individuals with extreme phenotypes by chance. For associations at the *CREBRF* locus (fasting glucose) and the *CNTN5* locus (LDL), the lead variants or variants in LD in Polynesians were either rare (frequency = 0.00-0.075) or not tested in sufficiently large AoU populations so we did not have the power to replicate these association using AoU. Therefore, the *CNTN5* association with LDL remains a potential novel population-enriched association (MAF = 0.023 in Samoans, 0.018 in Native Hawaiians) that awaits future validation.

## Discussion

In this work, we meta-analyzed Native Hawaiian and Samoan cohorts using imputed dosages from the latest TOPMed imputation server (release 3) for 13 cardiometabolic phenotypes. Despite improved imputation quality and increased sample size compared to previous studies, we do not observe any novel, replicable, commonly occurring Polynesian-enriched variants outside of the previously reported *CREBRF* and *CETP* missense variants.

Our analysis replicated 20 loci previously reported in the GWAS catalog (**Table 3**). Of the replicated loci, 10 passed the standard GWAS significance threshold of 5e-8 only through meta-analysis. Notably, we are able to detect signals for 5 previously reported Polynesian-enriched associations, *CREBRF* for BMI, hip circumference, waist circumference, and T2D (chr5:173108771, rs373863828) (14,15) as well as *CETP* for HDL (rs1597000001) (16). Despite being Polynesian-enriched alleles, these variants are well imputed with imputation Rsq of 0.991-0.994 and 0.957-1.0 for *CREBRF* and *CETP*, respectively. Though Polynesian-ancestry individuals are still not well represented and thus the reference panel is sub-optimal for imputing the genotypes of Polynesian-ancestry individuals, these two loci underscore the potential of the TOPMed imputation reference for identifying Polynesian-enriched association.

Through increased sample size in meta-analysis, our study was powered to test and confirm the association of *CREBRF* missense variant with fasting glucose (**Table 2**). This variant-phenotype association had previously been associated with type 2 diabetes (T2D) and nominally associated with fasting glucose levels in Samoans, Polynesians, and Pacific Islander cohorts (14,20,29,30). The non-replication of our top association (chr5:173045049, rs12513649) at the *CREBRF* locus in AoU, despite calculations showing sufficient power to replicate (Power = 99.62-100% at *p* = 0.01 level), is worth noting (**Supplementary Table 3**). Rs12513649 is found in AoU AMR and EAS populations at appreciable frequencies (0.0749 and 0.0661, respectively), but this is a variant in LD with the likely causal missense variant (rs373863828 (14)) only among Polynesian-ancestry individuals. Rs373863828 is extremely rare in AoU (AF of 0.00013, allele count of 109 of a possible 829,616) and does not have available summary statistics for comparison.

The remaining 4 novel variant-phenotype associations are relatively rare within our cohorts (AF 0.0017-0.023), with 3 of the 4 (*MYCBP2*, *BBS5*, *CNTN5*) commonly occurring in at least one global population (MAF > 1%; **Table 2**) and tested in AoU, but they fail to replicate. The relatively rare frequency of these variants within our cohort coincides with the lack of additional evidence in the association of variants in LD, possibly reflecting that these associations are technical artifacts. However, this does not rule out that these variants may instead capture the haplotype-tagging of unobserved associations (*e.g. CNTN5* in **Fig 2d**). In either case, improved imputation or sample size for Polynesian and related populations could help clarify whether these associations are due to an artifact of imputation or unobserved variants.

In our study, meta-analysis of Polynesian cohorts using the latest TOPMed imputation release serves to overcome some of the limitations of previous studies and explore whether more population-enriched associations are present. However, outside of the previously reported *CREBRF* and *CETP* missense variants, we found no additional Polynesian-enriched associations. Given the increased power of our study, our results informed the bounds of potential Polynesian-enriched associations yet to be discovered. It appears clear that both *CREBRF* and *CETP* are unique in terms of the combination of their allele frequency and effect sizes for the traits we examined, and they stand unmatched to other variants we were able to test (**Supplementary Fig 5**). While it is possible that other common Polynesian variants simply do not exert as strong of an effect on these traits, or that the environment has greater contribution to trait variation in these populations, we also note that current state-of-the-art imputation references still have room to improve for underrepresented populations such as the Polynesians and Pacific Islanders. Polynesian-enriched variants that are rare globally are more likely to be absent from the imputation reference or filtered out during the quality control process. Even for variants that can be found in the reference, because of inadequate haplotype representations, Pacific Islander populations are still relatively poorly imputed, particularly for Pacific Islanders with greater Polynesian ancestries compared to those with ancestries better represented in the references (31). This can be observed for the replicated locus on chromosome 19 around 45 Mb for LDL, triglycerides, and total cholesterol phenotypes: imputation accuracy in the admixed Native Hawaiians individuals (Rsq 0.95-1.00) is higher than the less unadmixed Samoan individuals (Rsq 0.66-0.71). Ergo, a natural future direction would be to use whole-genome sequencing (WGS) data for Polynesian-ancestry individuals to systematically discover Polynesian-enriched variation and create a representative reference panel that accurately imputes the genotype of potential Polynesian-specific variants. A recent study reported that a reference panel with as few as 24 unrelated WGS Samoan individuals could impute as many well-imputed variants (Rsq > 0.8) as the TOPMed panel for a Samoan cohort, and the performance exceeds that of TOPMed when 48 reference individuals were included (32). This highlights the large impact representation in reference panels may have for diverse populations. Such a reference panel can also be supplemented with other standard reference panels through meta-imputation (33), to reap the benefit of both the breadth of variation coverage across global populations and the depth of population-enriched variation.

In summary, we confirmed a metabolic trait’s association with the *CREBRF* locus in Polynesian populations, further supporting the pleiotropic nature of this locus (14–16). Although we did not identify other Polynesian-enriched associations for the traits we examined here, we note that there continue to be achievable advancements in available genomic resources, analytical strategies, and cohort aggregation that will allow a more comprehensive investigation to identify important genetic associations, whether for increasing or decreasing risk of complex diseases, or interacting with the environment, in these populations. Identification of these associations will not only reveal new biological insights but also reduce the disparity in how genomic medicine is practiced for these populations, such as the observed disparity in polygenic prediction models (34,35).

## Methods and Data

### MEC Native Hawaiian Cohort

The Multiethnic Cohort (MEC) Study is a prospective epidemiological cohort and consists of over 215,000 individuals from California and Hawai‘i. Height and weight were self-reported upon enrollment for most individuals with follow-up visits to collect biomarker data and questionnaires recording additional phenotypes such as waist and hip circumference. There are over 14,000 Native Hawaiians surveyed in the MEC with genetic data for 5,382 Native Hawaiian individuals available on 2 genotyping arrays, the Illumina multi-ethnic genotyping array (MEGA) and Infinium global diversity array (GDA). There are 4,144 individuals genotyped on MEGA with 1,093,693 variants available and 1,238 individuals genotyped on GDA with 1,549,554 variants available. Number of individuals available for each phenotype can be found in **Table 1**. Previously described QC protocol was followed prior to GRM inference (15,17,27,36). Some of the filtering criteria included minimum call rate, minor allele frequency, HWE p-value, poor clustering by visual inspection, mendelian errors, sex difference in allele frequencies for autosomes and sex chromosomes, and positional duplicates.

### Samoan Cohort

Participants for the Samoan cohort came from the Soifua Manuia (‘good health’) study, a population-based sample of 3,504 Samoan adults recruited in 2010 (37). Genotyping was performed for a subset of 3,182 Samoan adults using Genome-Wide Human SNP 6.0 arrays (Affymetrix) at 906,600 positions genome wide and extensive quality control was conducted. Height, weight, abdominal and hip circumferences were measured to the nearest 0.1 mm, 0.1 kg, and 1 mm, respectively, with participants in light island clothing. Measurements of total cholesterol, LDL- and HDL-cholesterol, triglycerides, glucose and insulin were derived from fasting serum samples following standard protocols. Additional details about phenotype measurement, sample genotyping, and genotype quality control have previously been described (14,38).

### Imputation to TOPMed Server

Native Hawaiians are available on two array platforms, MEGA and GDA, and each array was independently imputed to the TOPMed imputation server (release 3) to maximize the available variant information for each to the panel. Samoan array data was also imputed to TOPMed release 3. Before meta-analysis, variants were filtered to have a minimum imputed Rsq of 0.3 and minor allele count (MAC) > 0.

### Phenotype transformation

Previously studied phenotypes (all phenotypes with the exception of hip circumference, HOMA-IR, and waist circumference) followed previous published protocol accordingly (**Supplemental Table 4**) (39). Hip circumference, HOMA-IR, waist circumference phenotypes were transformed as follows:

HOMA-IR

fasting insulin (microU/L) x fasting glucose (nmol/L)/22.5
Hip circumference (cm)

sex stratified, regressed against age, and combined residuals
Waist circumference (cm)

sex stratified, regressed against age, and combined residuals

Covariates, transformations, and exclusion criteria were applied to each cohort independently. Individuals were stratified by sex before inverse normal transformations and combined after regressing on sex-specific covariates.

### Association testing with GENESIS

Both cohorts were analyzed using an in-house implementation of a previously reported GENESIS pipeline (40,41). For Native Hawaiians, inference of PCs and genetic relatedness matrix (GRM) used the intersection of variants between the GDA and MEGA arrays, approximately 1M variants.

This pipeline iteratively infers principal components (PCs) using PC-AiR and infers a GRM using PC-Relate. All programs and software were run with default settings and parameters. To conservatively account for potential structure, we include both the GRM and PCs in association testing when using GENESIS (20 PCs for Native Hawaiian and 3 PCs for Samoan associations).

### Meta-analysis with METAL

METAL (42) was used to meta-analyze the Native Hawaiian and Samoan summary stats using default parameters with the STDERR scheme. Prior to meta-analysis, variants with Rsq < 0.3 were removed from each cohort. Differences in variant ascertainment due to different array platforms may impact the imputation quality for some imputed variants. As a result, additional filtering criteria is added post analysis requiring variants to have imputed Rsq ≥ 0.3 in both cohorts and cross-cohort total MAC ≥ 10 to remove any very poorly or rare imputed associations from meta-analysis.

After meta-analysis, the top variants were examined against the GWAS catalog (26) for previously reported variants. Variants within 500kb of the top hit in a region were considered as part of the locus. Categorization using GWAS catalog is determined by the presence of a reported genome-wide significant hit within the locus (P < 5e-8); if such a hit was not present or if the reported hit was not genome-wide significant, the locus is considered newly confirmed. LocusZoom was used to plot the meta-analysis results at each locus with recombination rates (previously reported) and LD calculations (from this study using PLINK 1.90b7.2) for Native Hawaiians (27,43–45).

### Conditional analysis

Conditional analyses were run for novel associations and previously reported loci from the GWAS catalog. In both cases, imputed dosages for the variant being conditioned on were extracted from the imputed data, included as an additional fixed effect covariate, and association models were re-run.

We performed the following conditional analyses:

1. Potentially novel associations

a. The top signal in each association was conditioned on to detect potential secondary signals that could be masked by the primary one.
b. Phenotypes with associations at the *CREBRF* locus were conditioned on the known Polynesian-specific missense variant (rs373863828) to determine whether these associations may reflect the previously reported variant.
2. Associations with nearby reported signals

a. The dosage of the reported signal in GWAS catalog was conditioned on to verify whether the primary signal is reflecting it. In the case that there was more than one reported signal, conditional analysis was performed using the most significant variant from our meta-analysis.

### AoU Replication, Power Calculation

Due to the limited availability of external Polynesian genetic data, we attempted to replicate our potentially novel associations using the multi-ethnic samples from the All-of-Us Research Program for the same or related phenotypes. Gene-based and single variant associations for approximately 250,000 individuals were available on the All by All browser (https://allbyall.researchallofus.org). Summary statistics are available for 3,400 phenotypes. The lead variant at each locus was used when available; otherwise, a variant in linkage disequilibrium was used instead. We calculated power for each sub-cohort (AFR, AMR, EAS, EUR, and SAS) using the following R code (significance level for replication, alpha, was set at 0.01):

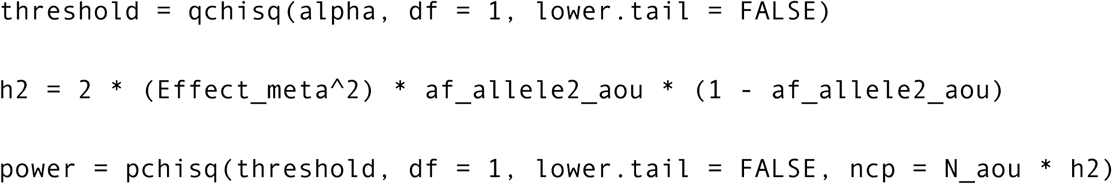

where Effect_meta is the effect size from the METAL meta-analysis, af_allele2_aou is the alternative allele frequency, and N_aou is the number of individuals.

### Artificial intelligence-based literature search

We followed a protocol described previously (46) to utilize large language models (LLMs, a specific subset of artificial intelligence) for the identification of causal genes at loci implicated via association studies. For each locus nominated in our study, we constructed a prompt including the genes and the phenotype in question and query ChatGPT (version 4o mini) to identify the likely causal gene and provide a confidence score between 0 and 1 (**Supplemental Text**). Overall, the confidence values for the selected genes were reasonably high for both novel loci and replicated loci (0.7-0.85 and 0.8-0.9, respectively). Across all responses for the 25 loci, we did not observe any hallucinations, nor did we observe responses conflicting with the given instructions. While the innerworkings of LLMs are opaque, the distribution of confidence scores would seem to reflect the expected amount of evidence existing in literature that would be available for the given categories, and likely comparable to a manual curation.

## Acknowledgements

We would like to thank all of the research participants in the Multiethnic Cohort that are involved in this study. The Multiethnic Cohort was funded through grants from the National Cancer Institute (U01CA164973, P01CA168530) and the National Human Genome Research Institute (U01HG007397). We would also like to thank the Samoan participants of the study, local village authorities, and the many Samoan and other field workers over the years. We acknowledge the Samoan Ministry of Health, the Samoa Bureau of Statistics, and the Samoan Ministry of Women, Community and Social Development for their support of this research. We give particular thanks to two research assistants, Melania Selu and Vaimoana Lupematasila, who contributed to the 2010 recruitment and continue to assist us in our work in Samoa. This work was funded by the National Institute of Health grants R01HL093093 (ST McGarvey), R01HL133040 (RL Minster), R01AG009375 (ST McGarvey), R01HL052611 (MI Kamboh), R01DK059642 (ST McGarvey), and R01DK055406 (R Deka). Genotyping for the Samoan cohort was performed in the Core Genotyping Laboratory at the University of Cincinnati, funded by National Institutes of Health grant P30ES006096 (S Pinney). We would like to acknowledge *All of Us* participants for their contributions and thank the National Institutes of Health’s All of Us Research Program for making their cohort analyses available. Analysis in this study was supported by grants from the National Human Genome Research Institute (R01HG011646 to C.W.K.C.), the National Institutes of Health (F31HG012159 to B.L.D.), and by the Center for Genetic Epidemiology First Year Graduate Fellowship (to X.W.). Computation for this work was supported by University of Southern California’s Center for Advanced Research Computing (https://www.carc.usc.edu/).

## Data Availability

The analysis pipeline for the Samoan cohort using GENESIS can be found at: https://github.com/UW-GAC/analysis_pipeline. This pipeline was adapted to run on USC’s Center for Advanced Research Computing (CARC) systems for the Native Hawaiians and can be found at: https://github.com/bldinh/carc_topmed_analysis_pipeline.

Data for The Native Hawaiian and Samoan cohorts are available on dbGaP (study accessions: phs002183.v2.p1 and phs000914.v1.p1); data access will require the collaboration letter with the Native Hawaiian Community Advisory Board and the Samoan Health Research Council, respectively.

## Notes

### Competing Interest Statement

The authors have declared no competing interest.

### Author Declarations

The ethnics committee/IRB of University of Southern California gave ethical approval for this work.

## References

1. Aluli NE, Jones KL, Reyes PW, Brady SK, Tsark JU, Howard BV. Diabetes and Cardiovascular Risk Factors in Native Hawaiians. Hawaii Med J. 2009 Aug;68(7):152–7. PubMed PMID: 19653416; PubMed Central PMCID: PMC4381427.

2. Đoàn LN, Takata Y, Hooker K, Mendez-Luck C, Irvin VL. Trends in Cardiovascular Disease by Asian American, Native Hawaiian, and Pacific Islander Ethnicity, Medicare Health Outcomes Survey 2011–2015. J Gerontol A Biol Sci Med Sci. 2022 Feb 1;77(2):299–309. doi:10.1093/gerona/glab262

3. Mau MK, Sinclair K, Saito EP, Baumhofer KN, Kaholokula JK. Cardiometabolic Health Disparities in Native Hawaiians and Other Pacific Islanders. Epidemiol Rev. 2009 Nov 1;31(1):113–29. doi:10.1093/ajerev/mxp004

4. Tung WC, Barnes M. Heart Diseases Among Native Hawaiians and Pacific Islanders. Home Health Care Management & Practice. 2014 May 1;26(2):110–3. doi:10.1177/1084822313516125

5. Hawley NL, Minster RL, Weeks DE, Viali S, Reupena MS, Sun G, et al. Prevalence of adiposity and associated cardiometabolic risk factors in the samoan genome-wide association study. American Journal of Human Biology. 2014;26(4):491–501. doi:10.1002/ajhb.22553

6. Lin S, Naseri T, Linhart C, Morrell S, Taylor R, McGarvey ST, et al. Trends in diabetes and obesity in Samoa over 35 years, 1978–2013. Diabetic Medicine. 2017;34(5):654–61. doi:10.1111/dme.13197

7. Ha EK, Shriner D, Callier SL, Riley L, Adeyemo AA, Rotimi CN, et al. Native Hawaiian and Pacific Islander populations in genomic research. npj Genomic Medicine. 2024 Sep 30;9(1):45. doi:10.1038/s41525-024-00428-6

8. Moore MA, Baumann F, Foliaki S, Goodman MT, Haddock R, Maraka R, et al. Cancer Epidemiology in the Pacific Islands-Past, Present and Future. Asian Pac J Cancer Prev. 2010;11(0 2):99–106. PubMed PMID: 20553071; PubMed Central PMCID: PMC4386924.

9. Haque AT, Berrington de González A, Chen Y, Haozous EA, Inoue-Choi M, Lawrence WR, et al. Cancer mortality rates by racial and ethnic groups in the United States, 2018-2020. J Natl Cancer Inst. 2023 Jul 1;115(7):822–30. doi:10.1093/jnci/djad069

10. Chiang CWK. The Opportunities and Challenges of Integrating Population Histories Into Genetic Studies for Diverse Populations: A Motivating Example From Native Hawaiians. Front Genet. 2021 Sep 27;12:643883. doi:10.3389/fgene.2021.643883 PubMed PMID: 34646295; PubMed Central PMCID: PMC8503554.

11. Popejoy AB, Fullerton SM. Genomics is failing on diversity. Nature News. 2016 Oct 13;538(7624):161. doi:10.1038/538161a

12. Fatumo S, Chikowore T, Choudhury A, Ayub M, Martin AR, Kuchenbaecker K. A roadmap to increase diversity in genomic studies. Nature Medicine. 2022 Feb;28(2):243–50. doi:10.1038/s41591-021-01672-4

13. Emde AK, Phipps-Green A, Cadzow M, Gallagher CS, Major TJ, Merriman ME, et al. Mid-pass whole genome sequencing enables biomedical genetic studies of diverse populations. BMC Genomics. 2021 Nov 1;22(1):666. doi:10.1186/s12864-021-07949-9

14. Minster RL, Hawley NL, Su CT, Sun G, Kershaw EE, Cheng H, et al. A thrifty variant in CREBRF strongly influences body mass index in Samoans. Nature Genetics. 2016 Sep;48(9):9. doi:10.1038/ng.3620

15. Lin M, Caberto C, Wan P, Li Y, Lum-Jones A, Tiirikainen M, et al. Population-specific reference panels are crucial for genetic analyses: an example of the CREBRF locus in Native Hawaiians. Hum Mol Genet. 2020 Aug 3;29(13):2275–84. doi:10.1093/hmg/ddaa083 PubMed PMID: 32491157; PubMed Central PMCID: PMC7399533.

16. Moors J, Krishnan M, Sumpter N, Takei R, Bixley M, Cadzow M, et al. A Polynesian-specific missense CETP variant alters the lipid profile. HGG Adv. 2023 May 8;4(3):100204. doi:10.1016/j.xhgg.2023.100204 PubMed PMID: 37250494; PubMed Central PMCID: PMC10209881.

17. Sun H, Lin M, Russell EM, Minster RL, Chan TF, Dinh BL, et al. The impact of global and local Polynesian genetic ancestry on complex traits in Native Hawaiians. PLOS Genetics. 2021 Feb 11;17(2):e1009273. doi:10.1371/journal.pgen.1009273

18. Harris DN, Kessler MD, Shetty AC, Weeks DE, Minster RL, Browning S, et al. Evolutionary history of modern Samoans. Proceedings of the National Academy of Sciences. 2020 Apr 28;117(17):9458–65. doi:10.1073/pnas.1913157117

19. Naka I, Furusawa T, Kimura R, Natsuhara K, Yamauchi T, Nakazawa M, et al. A missense variant, rs373863828-A (p.Arg457Gln), of CREBRF and body mass index in Oceanic populations. J Hum Genet. 2017 Sep;62(9):847–9. doi:10.1038/jhg.2017.44

20. Hanson RL, Safabakhsh S, Curtis JM, Hsueh WC, Jones LI, Aflague TF, et al. Association of CREBRF variants with obesity and diabetes in Pacific Islanders from Guam and Saipan. Diabetologia. 2019 Sep 1;62(9):1647–52. doi:10.1007/s00125-019-4932-z

21. Berry SD, Walker CG, Ly K, Snell RG, Atatoa Carr PE, Bandara D, et al. Widespread prevalence of a CREBRF variant amongst Māori and Pacific children is associated with weight and height in early childhood. Int J Obes. 2018 Apr;42(4):603–7. doi:10.1038/ijo.2017.230

22. Zhang JZ, Heinsberg LW, Krishnan M, Hawley NL, Major TJ, Carlson JC, et al. Multivariate analysis of a missense variant in CREBRF reveals associations with measures of adiposity in people of Polynesian ancestries. Genetic Epidemiology. 2023;47(1):105–18. doi:10.1002/gepi.22508

23. Chen S, Francioli LC, Goodrich JK, Collins RL, Kanai M, Wang Q, et al. A genome-wide mutational constraint map quantified from variation in 76,156 human genomes [Internet]. bioRxiv; 2022 [cited 2023 Nov 8]. p. 2022.03.20.485034. Available from: https://www.biorxiv.org/content/10.1101/2022.03.20.485034v2 doi:10.1101/2022.03.20.485034

24. Taliun D, Harris DN, Kessler MD, Carlson J, Szpiech ZA, Torres R, et al. Sequencing of 53,831 diverse genomes from the NHLBI TOPMed Program. Nature. 2021 Feb;590(7845):290–9. doi:10.1038/s41586-021-03205-y

25. TOPMed Imputation Server [Internet]. [cited 2025 Oct 29]. Available from: https://imputation.biodatacatalyst.nhlbi.nih.gov/#!pages/about

26. Cerezo M, Sollis E, Ji Y, Lewis E, Abid A, Bircan KO, et al. The NHGRI-EBI GWAS Catalog: standards for reusability, sustainability and diversity. Nucleic Acids Res. 2025 Jan 6;53(D1):D998–1005. doi:10.1093/nar/gkae1070

27. Dinh BL, Tang E, Taparra K, Nakatsuka N, Chen F, Chiang CWK. Recombination map tailored to Native Hawaiians may improve robustness of genomic scans for positive selection. Hum Genet. 2024;143(1):85–99. doi:10.1007/s00439-023-02625-2 PubMed PMID: 38157018; PubMed Central PMCID: PMC10794367.

28. null null. The “All of Us” Research Program. New England Journal of Medicine. 2019 Aug 15;381(7):668–76. doi:10.1056/NEJMsr1809937

29. Krishnan M, Major TJ, Topless RK, Dewes O, Yu L, Thompson JMD, et al. Discordant association of the CREBRF rs373863828 A allele with increased BMI and protection from type 2 diabetes in Māori and Pacific (Polynesian) people living in Aotearoa/New Zealand. Diabetologia. 2018 Jul 1;61(7):1603–13. doi:10.1007/s00125-018-4623-1

30. Russell EM, Carlson JC, Krishnan M, Hawley NL, Sun G, Cheng H, et al. CREBRF missense variant rs373863828 has both direct and indirect effects on type 2 diabetes and fasting glucose in Polynesian peoples living in Samoa and Aotearoa New Zealand. BMJ Open Diab Res Care. 2022 Feb 10;10(1). doi:10.1136/bmjdrc-2021-002275 PubMed PMID: 10.1136/bmjdrc-2021-002275.

31. Cahoon JL, Rui X, Tang E, Simons C, Langie J, Chen M, et al. Imputation Efficacy Across Global Human Populations [Internet]. bioRxiv; 2023 [cited 2023 Oct 27]. p. 2023.05.22.541241. Available from: https://www.biorxiv.org/content/10.1101/2023.05.22.541241v1 doi:10.1101/2023.05.22.541241

32. Carlson JC, Krishnan M, Liu S, Anderson KJ, Zhang JZ, Spor LM, et al. The Soifua Manuia reference panel with 2,570 Samoan haplotypes improves genotype imputation quality among Samoans. medRxiv. 2025 Apr 30;2023.10.31.23297835. doi:10.1101/2023.10.31.23297835 PubMed PMID: 37961708; PubMed Central PMCID: PMC10635250.

33. Yu K, Das S, LeFaive J, Kwong A, Pleiness J, Forer L, et al. Meta-imputation: An efficient method to combine genotype data after imputation with multiple reference panels. The American Journal of Human Genetics. 2022 Jun 2;109(6):1007–15. doi:10.1016/j.ajhg.2022.04.002

34. Lo YC, Tian H, Chan TF, Jeon S, Alatorre K, Dinh BL, et al. The accuracy of polygenic score models for BMI and Type II diabetes in the Native Hawaiian population. Commun Biol. 2025 Apr 23;8(1):651. doi:10.1038/s42003-025-08050-7

35. Lee EY, Dinh BL, Tang J, Streicher S, Wang X, Biswas S, et al. Incorporating Dietary Information to Enhance Polygenic Prediction Models with Applications to Body Mass Index and Type 2 Diabetes [Internet]. medRxiv; 2025 [cited 2026 Jan 22]. p. 2025.10.03.25337287. Available from: https://www.medrxiv.org/content/10.1101/2025.10.03.25337287v2 doi:10.1101/2025.10.03.25337287

36. Bogumil D, Sheng X, Wan P, Xia L, Pooler L, Cheng I, et al. The Multiethnic Cohort: A Resource for the study of Genetic and non-Genetic Cancer Risk Across Populations [Internet]. medRxiv; 2025 [cited 2025 Aug 7]. p. 2025.06.09.25328993. Available from: https://www.medrxiv.org/content/10.1101/2025.06.09.25328993v1 doi:10.1101/2025.06.09.25328993

37. Hawley NL, Minster RL, Weeks DE, Viali S, Reupena MS, Sun G, et al. Prevalence of Adiposity and Associated Cardiometabolic Risk Factors in the Samoan Genome-Wide Association Study. Am J Hum Biol. 2014;26(4):491–501. doi:10.1002/ajhb.22553 PubMed PMID: 24799123; PubMed Central PMCID: PMC4292846.

38. Hawley NL, Minster RL, Weeks DE, Viali S, Reupena MS, Sun G, et al. Prevalence of adiposity and associated cardiometabolic risk factors in the samoan genome-wide association study. American Journal of Human Biology. 2014;26(4):491–501. doi:10.1002/ajhb.22553

39. Wojcik GL, Graff M, Nishimura KK, Tao R, Haessler J, Gignoux CR, et al. Genetic analyses of diverse populations improves discovery for complex traits. Nature. 2019 Jun;570(7762):514–8. doi:10.1038/s41586-019-1310-4 PubMed PMID: 31217584; PubMed Central PMCID: PMC6785182.

40. UW-GAC/analysis_pipeline [R] [Internet]. GAC; 2025 [cited 2025 Oct 29]. Available from: https://github.com/UW-GAC/analysis_pipeline

41. Gogarten SM, Sofer T, Chen H, Yu C, Brody JA, Thornton TA, et al. Genetic association testing using the GENESIS R/Bioconductor package. Bioinformatics. 2019 Dec 15;35(24):5346–8. doi:10.1093/bioinformatics/btz567

42. Willer CJ, Li Y, Abecasis GR. METAL: fast and efficient meta-analysis of genomewide association scans. Bioinformatics. 2010 Sep 1;26(17):2190–1. doi:10.1093/bioinformatics/btq340 PubMed PMID: 20616382; PubMed Central PMCID: PMC2922887.

43. Pruim RJ, Welch RP, Sanna S, Teslovich TM, Chines PS, Gliedt TP, et al. LocusZoom: regional visualization of genome-wide association scan results. Bioinformatics. 2010 Sep 15;26(18):2336–7. doi:10.1093/bioinformatics/btq419

44. Purcell S, Chang CC. PLINK 1.9 [Internet]. Available from: https://www.cog-genomics.org/plink/1.9/

45. Chang CC, Chow CC, Tellier LC, Vattikuti S, Purcell SM, Lee JJ. Second-generation PLINK: rising to the challenge of larger and richer datasets. Gigascience. 2015 Dec 1;4(1):s13742-015-0047–8. doi:10.1186/s13742-015-0047-8

46. Shringarpure SS, Wang W, Karagounis S, Wang X, Reisetter AC, Auton A, et al. Large language models identify causal genes in complex trait GWAS [Internet]. medRxiv; 2024 [cited 2024 Dec 19]. p. 2024.05.30.24308179. Available from: https://www.medrxiv.org/content/10.1101/2024.05.30.24308179v1 doi:10.1101/2024.05.30.24308179

