## Supplemental Figures and Tables for "Meta-analysis of over 8,000 individuals from Hawai‘i and Samoa for genetic associations to cardiometabolic phenotypes"

### Supplementary Figures

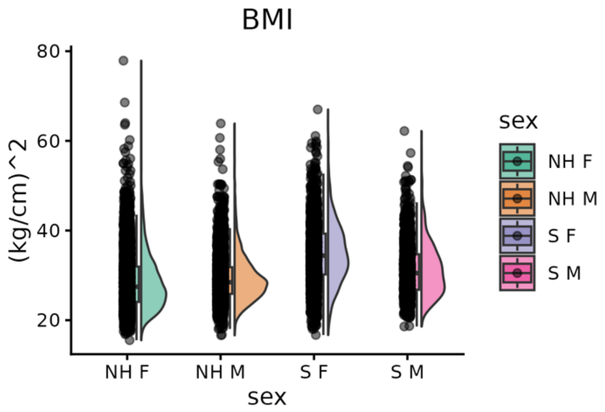


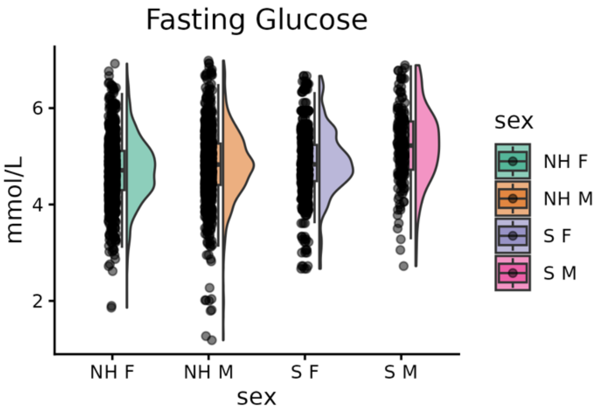


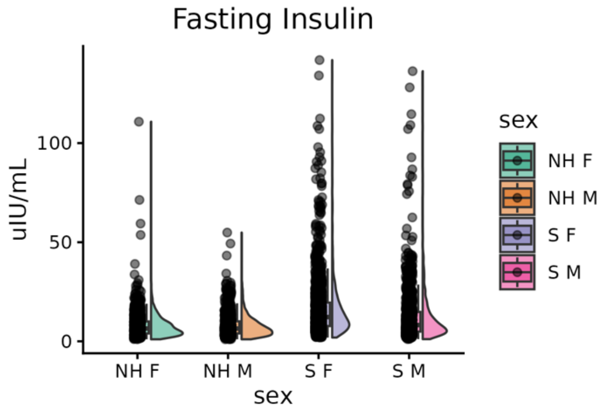


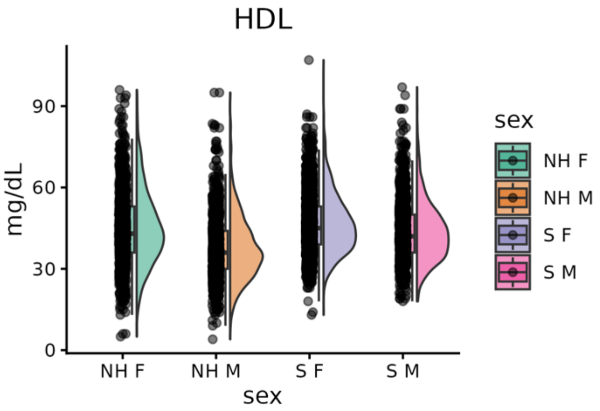


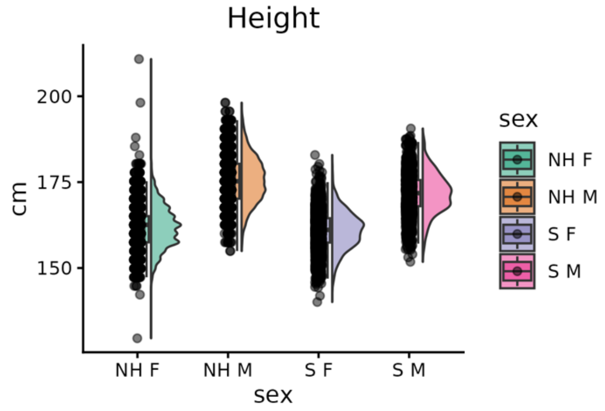


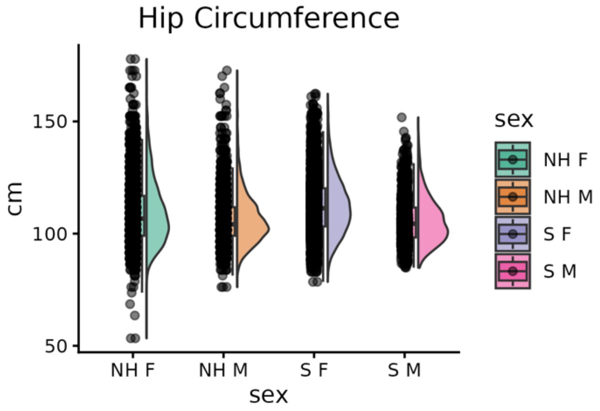

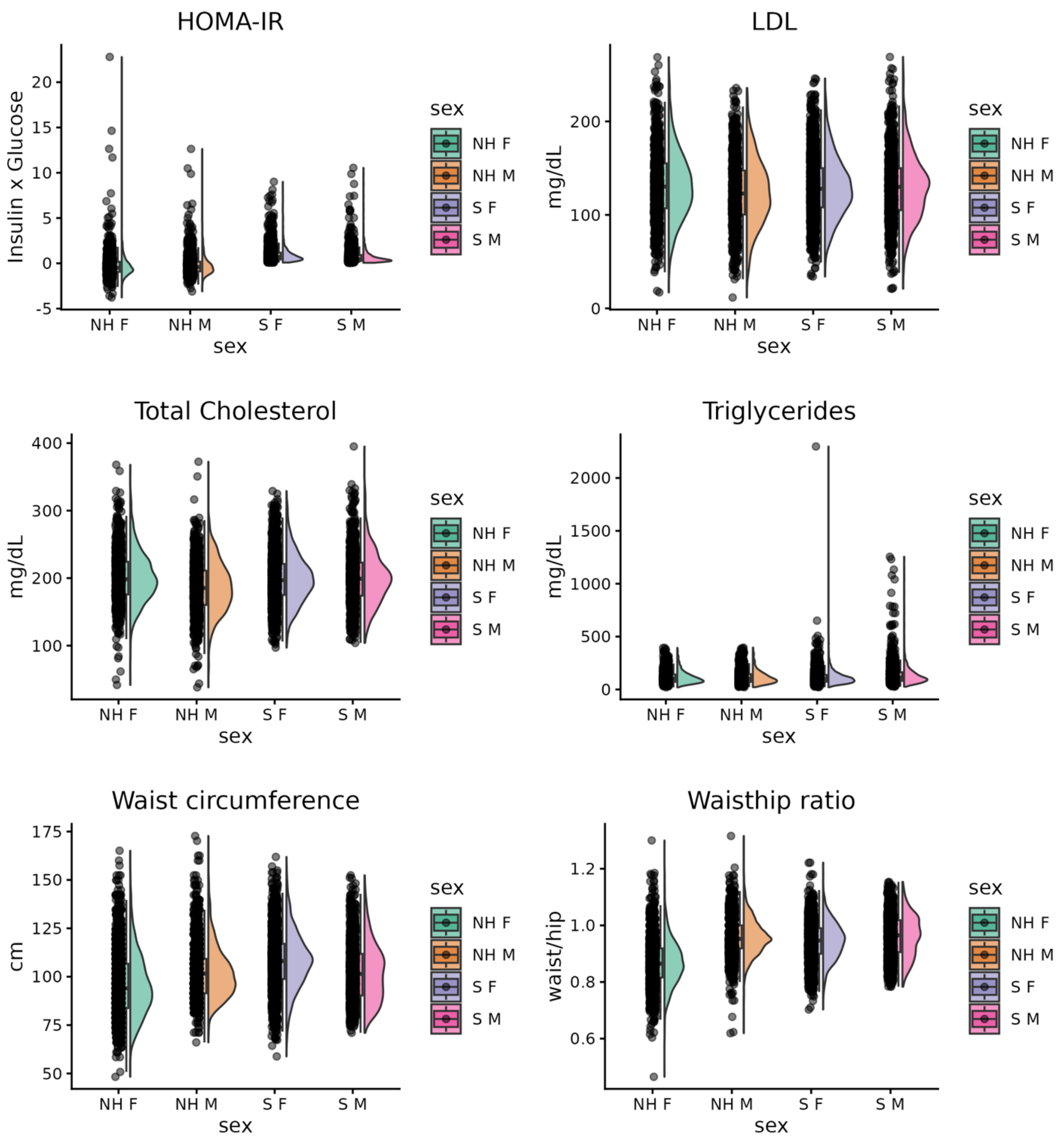


**Supplementary Figure 1.** Phenotype distributions for cohorts before unit harmonization and normalization. T2D is the only binary trait of the studied phenotypes. Native Hawaiians (N=5343): 2564 cases, 2779 controls. Samoans (N=2875): XXX cases, YYY controls.

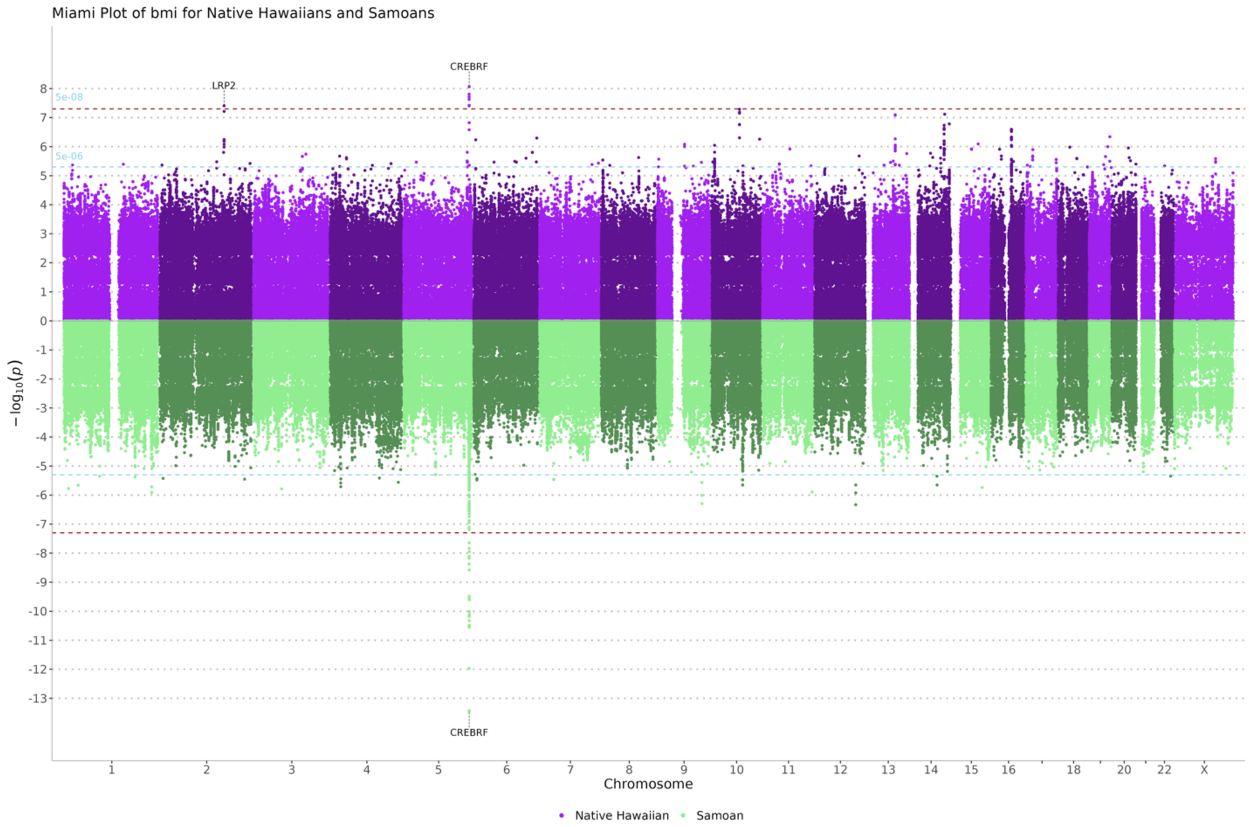
Miami Plot of bmi for Native Hawaiians and Samoans
CREBRF
LRP2
5e-08
5e-06
CREBRF
$-\log_{10}(p)$
Chromosome
1 2 3 4 5 6 7 8 9 10 11 12 13 14 15 16 18 20 22 X
Native Hawaiian
Samoan

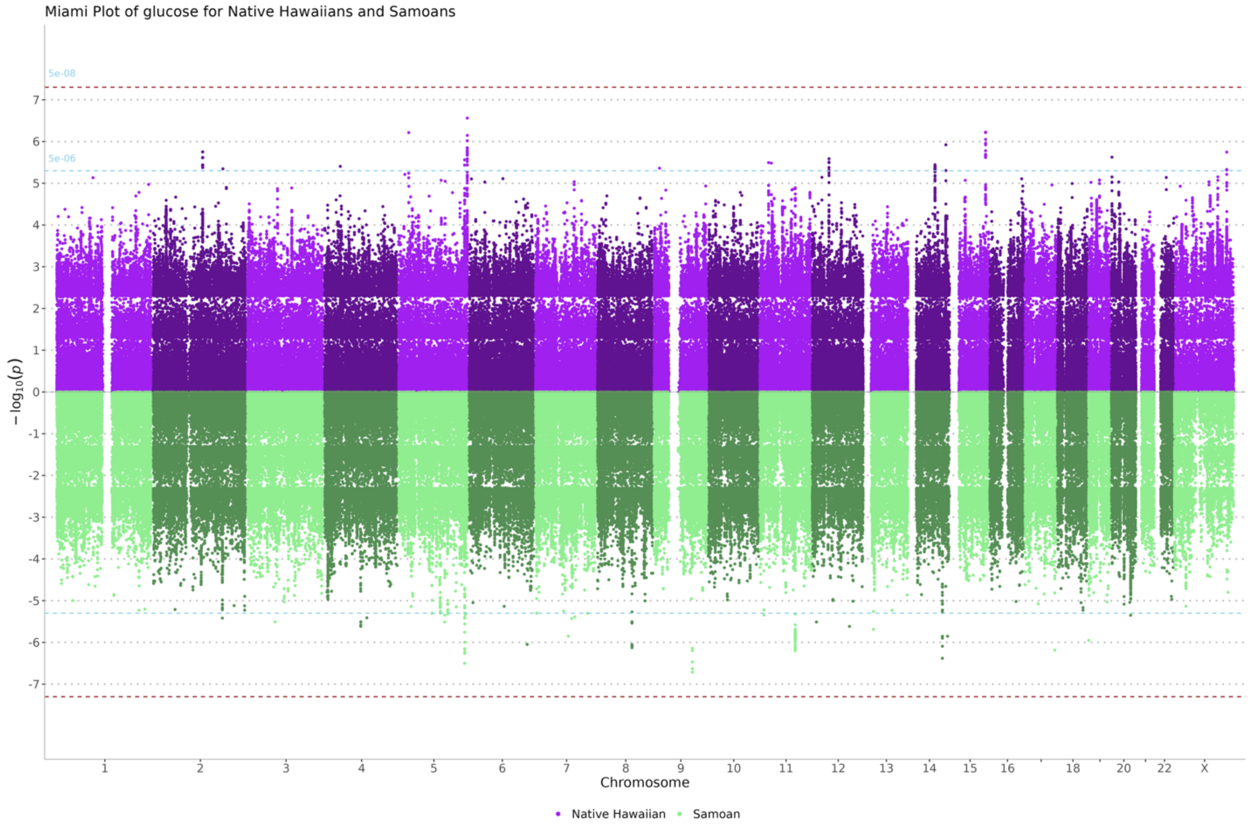
Miami Plot of glucose for Native Hawaiians and Samoans
5e-08
5e-06
$-\log_{10}(p)$
Chromosome
1 2 3 4 5 6 7 8 9 10 11 12 13 14 15 16 18 20 22 X
Native Hawaiian
Samoan

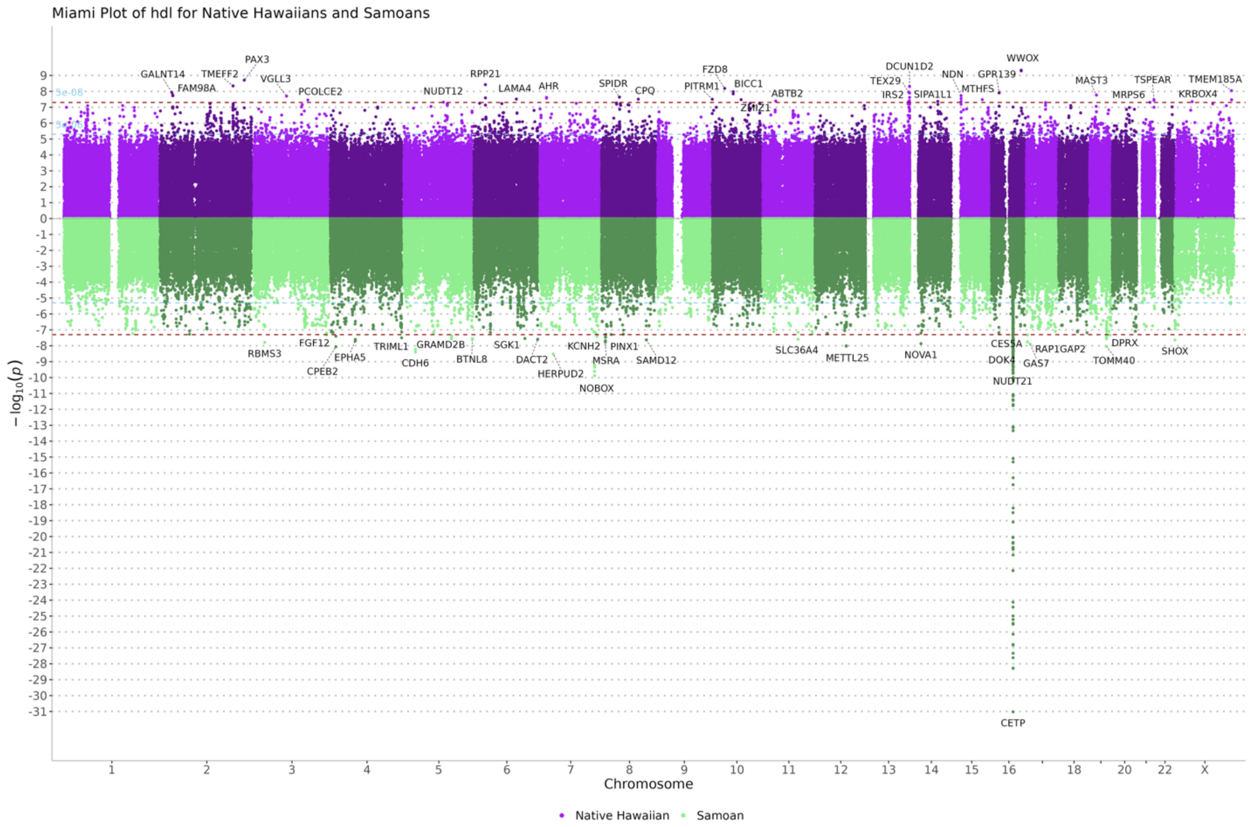

Miami Plot of hdl for Native Hawaiians and Samoans
$-\log_{10}(p)$
Chromosome
Native Hawaiian
Samoan
GALNT14
FAM98A
TMEFF2
PAX3
VGLL3
PCOLCE2
NUDT12
RPP21
LAMA4
AHR
SPIDR
CPQ
PITRM1
FZD8
BICC1
ZMIZ1
ABTB2
TEX29
DCUN1D2
IRS2
SIPA1L1
NDN
MTHFS
GPR139
WWOX
MAST3
MRPS6
TSPEAR
TMEM185A
KRBOX4
RBMS3
FGF12
CPEB2
EPHA5
TRIML1
CDH6
GRAMD2B
BTNL8
SGK1
DACT2
HERPUD2
KCNH2
NOBOX
MSRA
PINX1
SAMD12
SLC36A4
METTL25
NOVA1
CES5A
DOK4
NUDT21
GAS7
RAP1GAP2
TOMM40
DPRX
SHOX
CETP


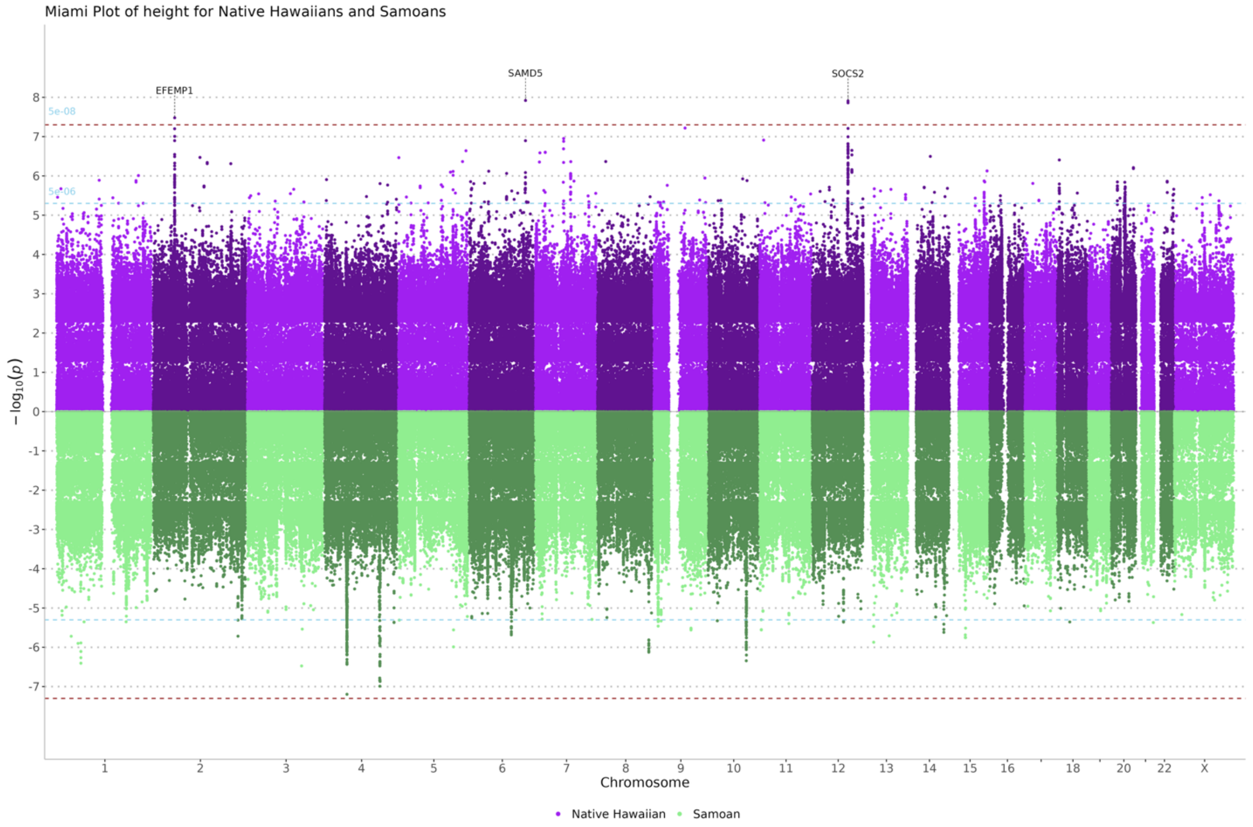

Miami Plot of height for Native Hawaiians and Samoans
$-\log_{10}(p)$
Chromosome
Native Hawaiian
Samoan
5e-08
5e-06
EFEMP1
SAMD5
SOCS2

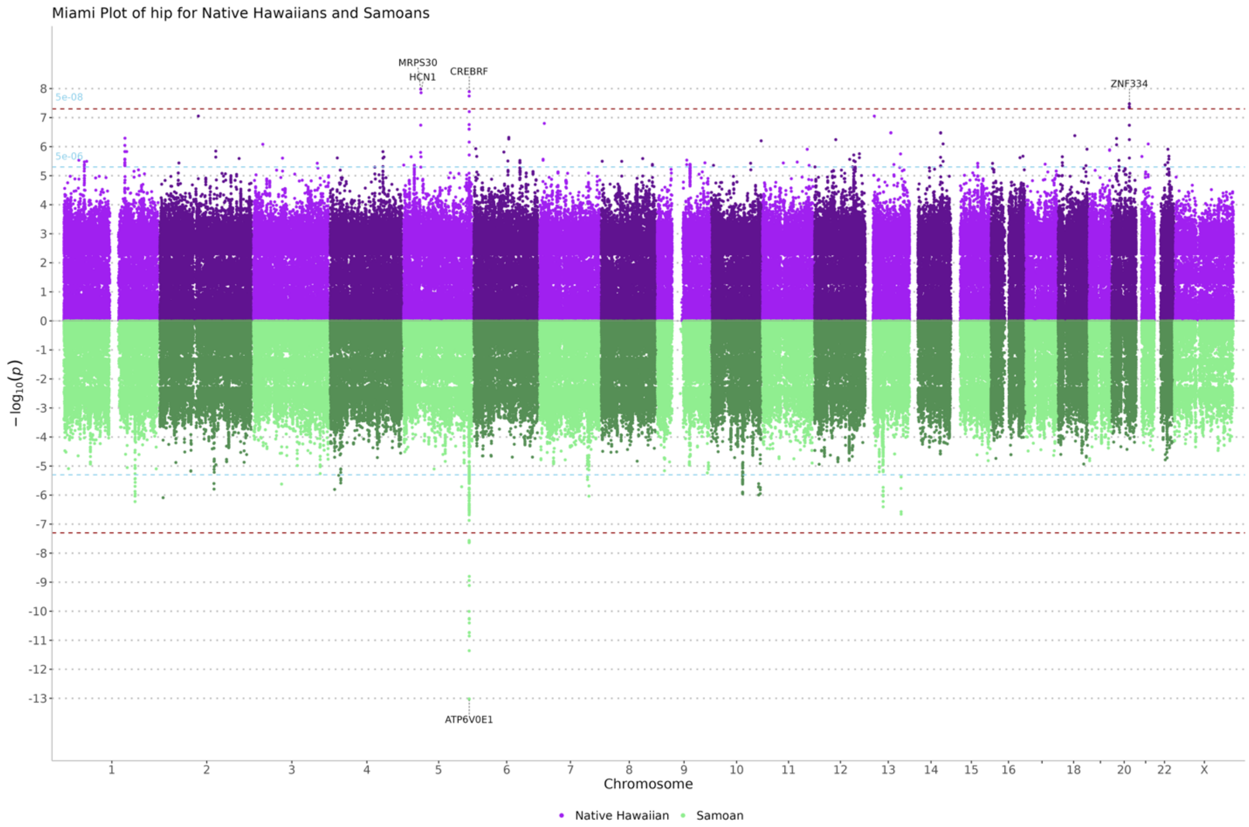

Miami Plot of hip for Native Hawaiians and Samoans
MRPS30
HCN1
CREBRF
ZNF334
ATP6V0E1
5e-08
5e-06
$-\log_{10}(p)$
Chromosome
Native Hawaiian
Samoan


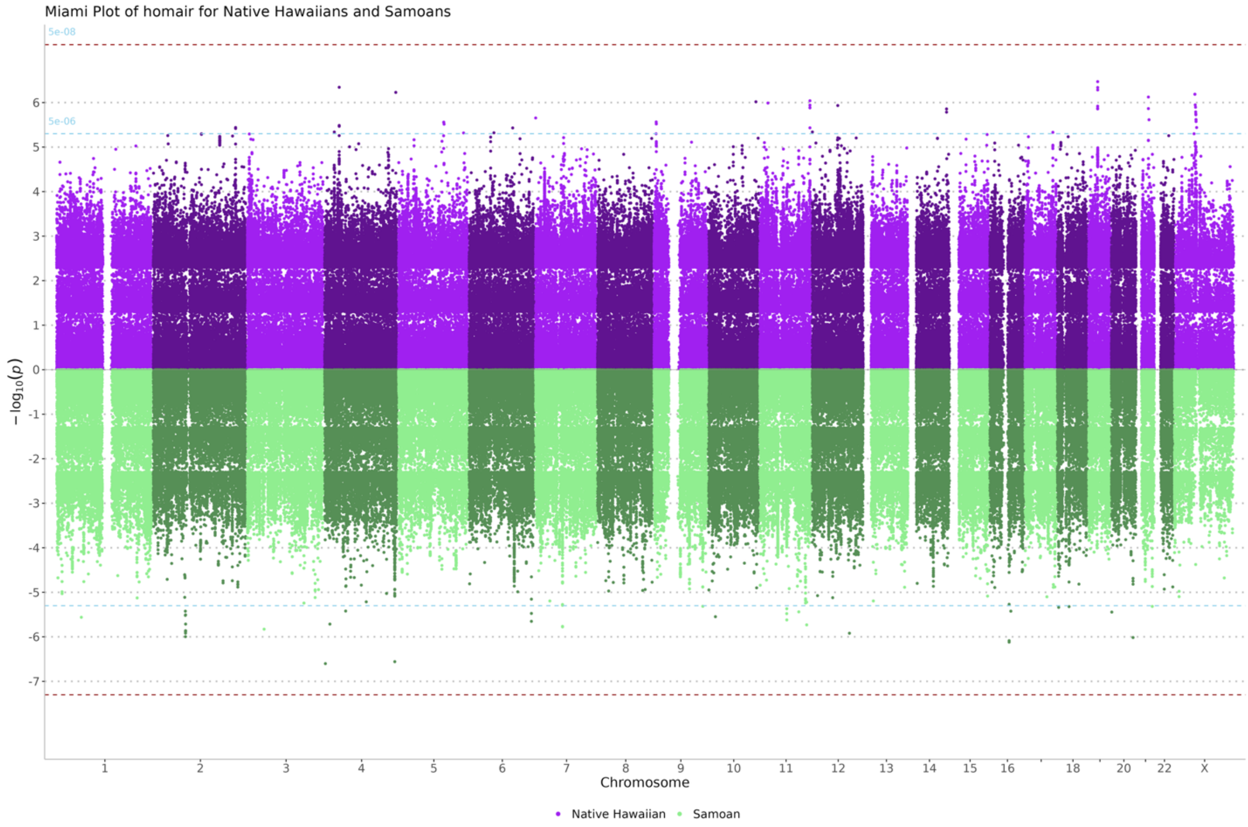

Miami Plot of homair for Native Hawaiians and Samoans
5e-08
5e-06
$-\log_{10}(p)$
Chromosome
Native Hawaiian
Samoan

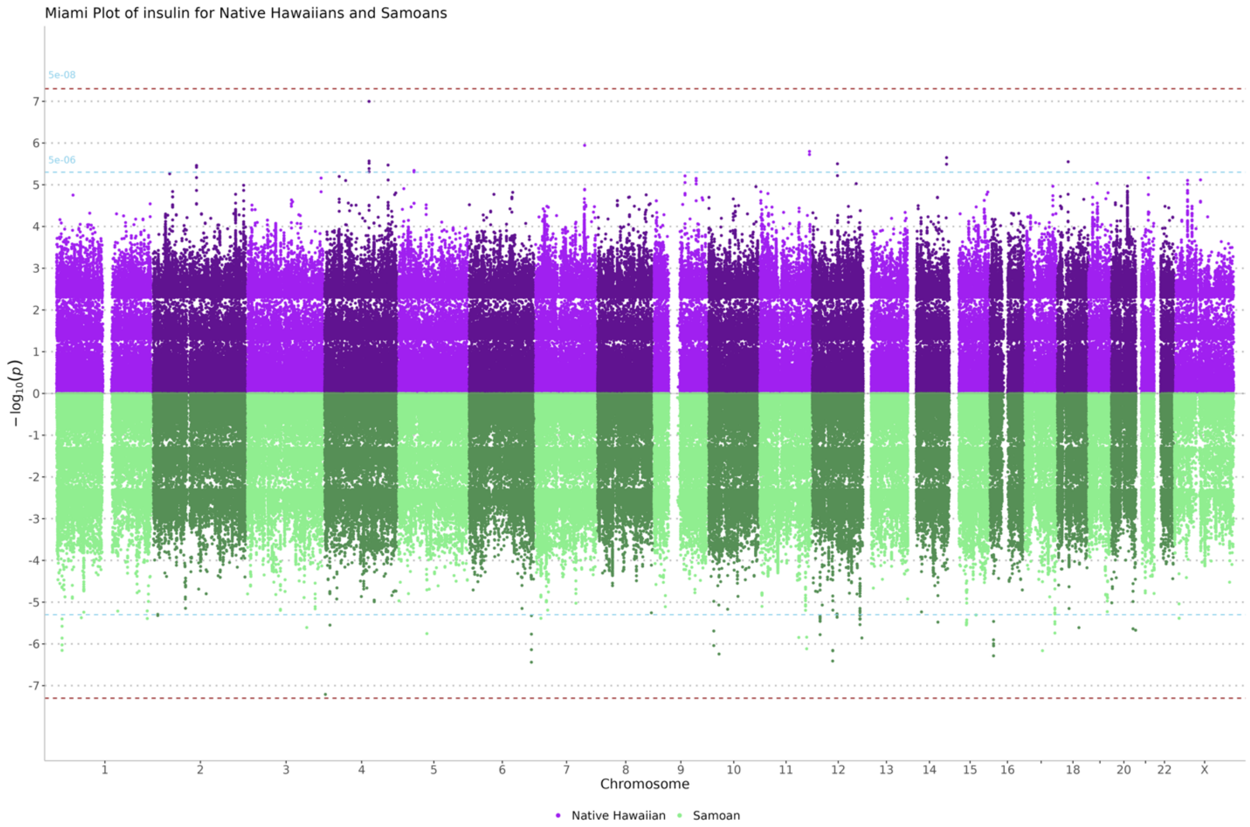

Miami Plot of insulin for Native Hawaiians and Samoans
5e-08
5e-06
$-\log_{10}(p)$
Chromosome
Native Hawaiian
Samoan


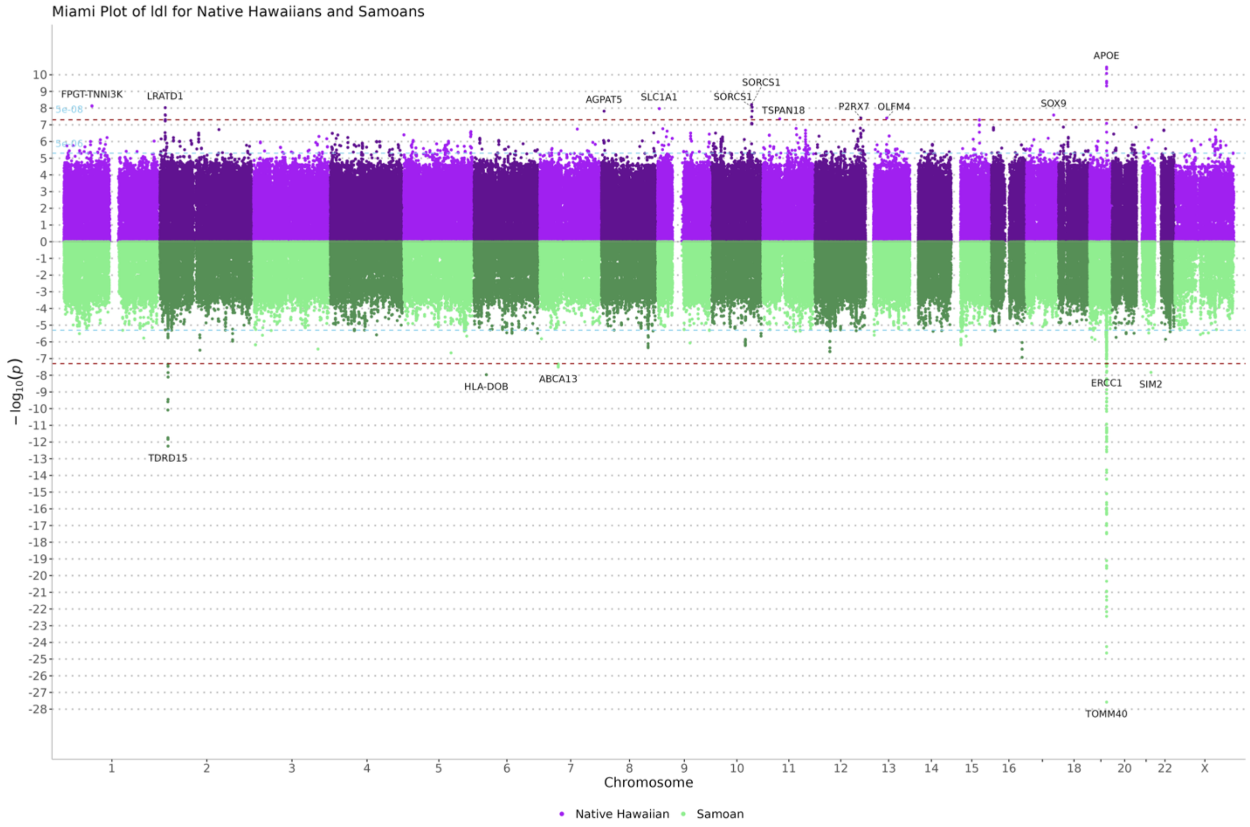

Miami Plot of ldl for Native Hawaiians and Samoans
APOE
FPGT-TNNI3K
LRATD1
AGPAT5
SLC1A1
SORCS1
SORCS1
TSPAN18
P2RX7
OLFM4
SOX9
HLA-DOB
ABCA13
ERCC1
SIM2
TDRD15
TOMM40
$-\log_{10}(p)$
Chromosome
Native Hawaiian
Samoan

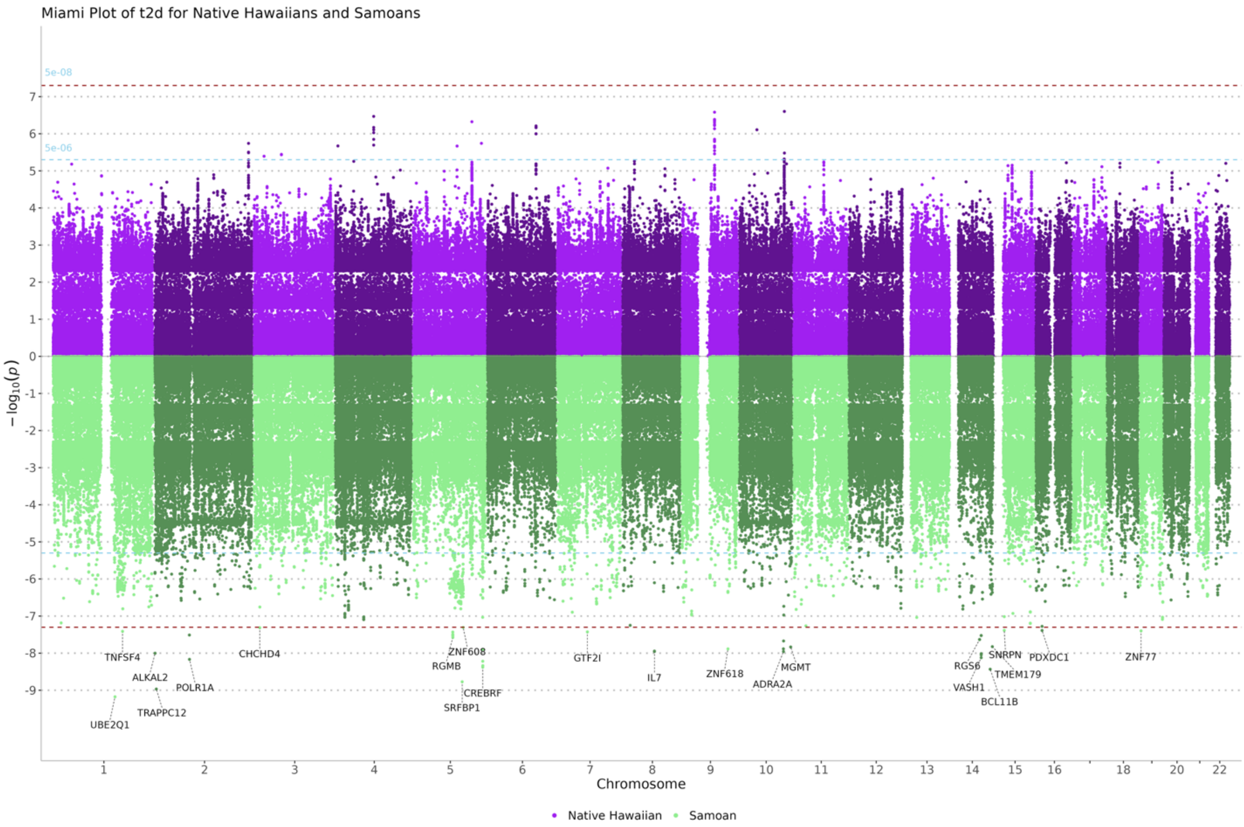
Miami Plot of t2d for Native Hawaiians and Samoans
5e-08
5e-06
$-\log_{10}(p)$
Chromosome
Native Hawaiian
Samoan
TNFSF4
ALKAL2
POLR1A
TRAPPC12
UBE2Q1
CHCHD4
ZNF608
RGMB
CREBRF
SRFBP1
GTF2I
IL7
ZNF618
MGMT
ADRA2A
RGS6
VASH1
SNRPN
TMEM179
BCL11B
PDXDC1
ZNF77

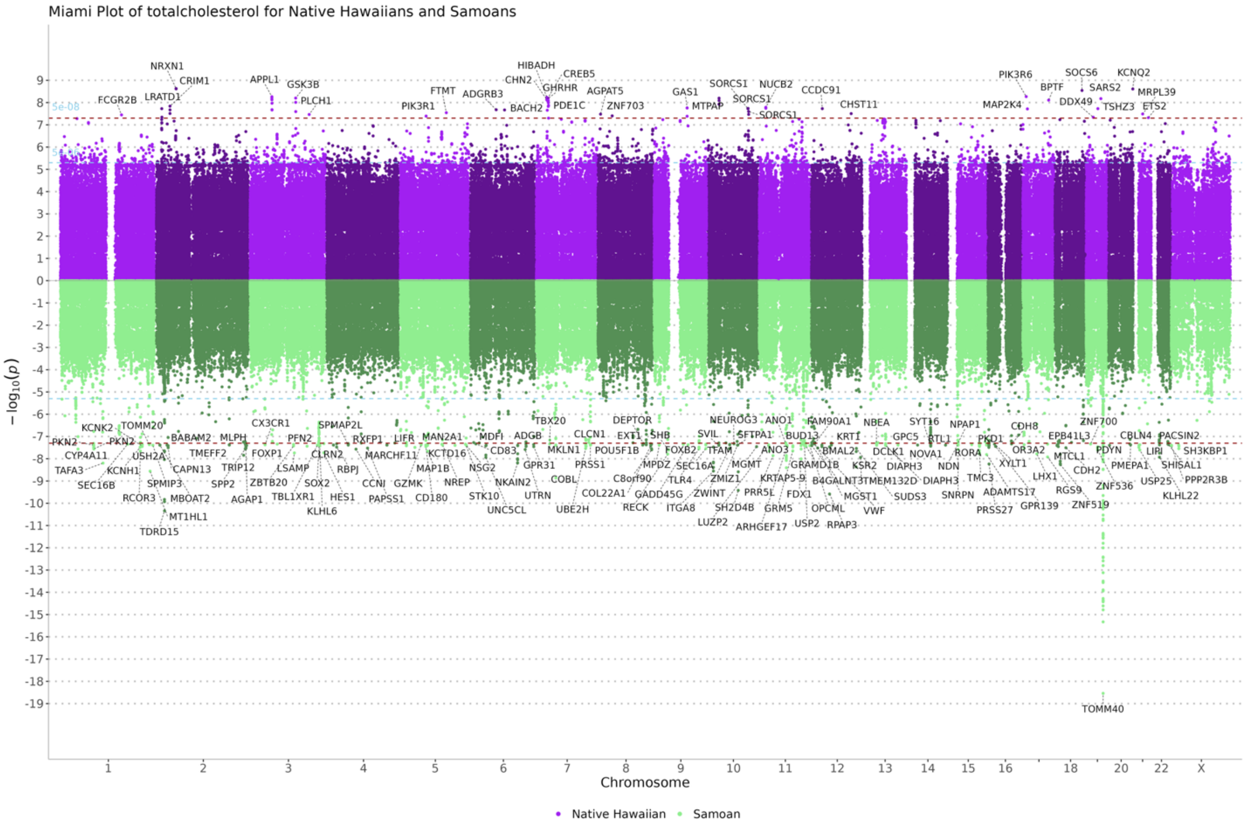
Miami Plot of totalcholesterol for Native Hawaiians and Samoans
5e-08
$-\log_{10}(p)$
Chromosome
Native Hawaiian
Samoan
NRXN1
CRIM1
LRATD1
FCGR2B
APPL1
GSK3B
PLCH1
PIK3R1
FTMT
ADGRB3
HIBADH
CHN2
CREB5
GHRHR
PDE1C
BACH2
AGPAT5
ZNF703
GAS1
MTPAP
SORCS1
NUCB2
CCDC91
CHST11
PIK3R6
MAP2K4
SOCS6
BPTF
SARS2
DDX49
KCNQ2
MRPL39
TSHZ3
ETS2
TOMM40

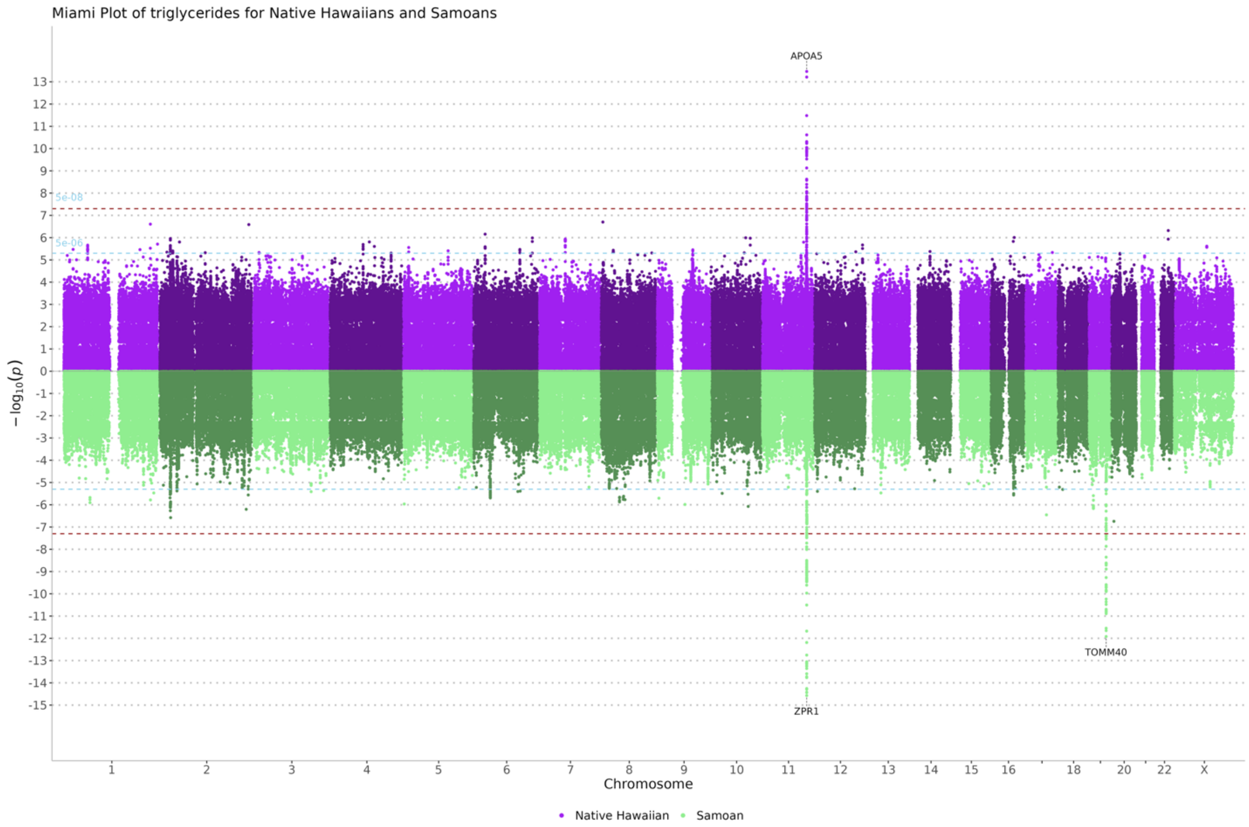
Miami Plot of triglycerides for Native Hawaiians and Samoans
APOA5
ZPR1
TOMM40
5e-08
5e-06
$-\log_{10}(p)$
Chromosome
Native Hawaiian
Samoan

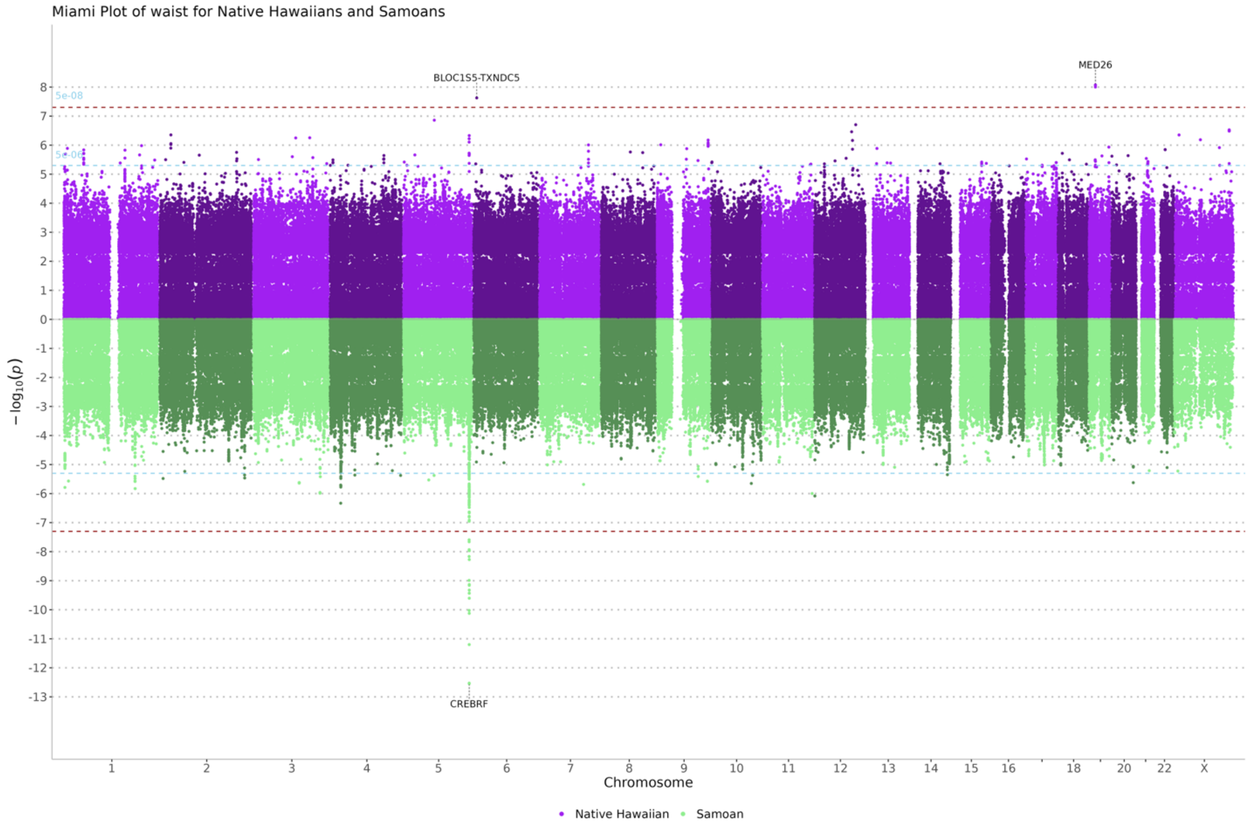
Miami Plot of waist for Native Hawaiians and Samoans
BLOC1S5-TXNDC5
MED26
CREBRF
5e-08
$-\log_{10}(p)$
Chromosome
Native Hawaiian
Samoan

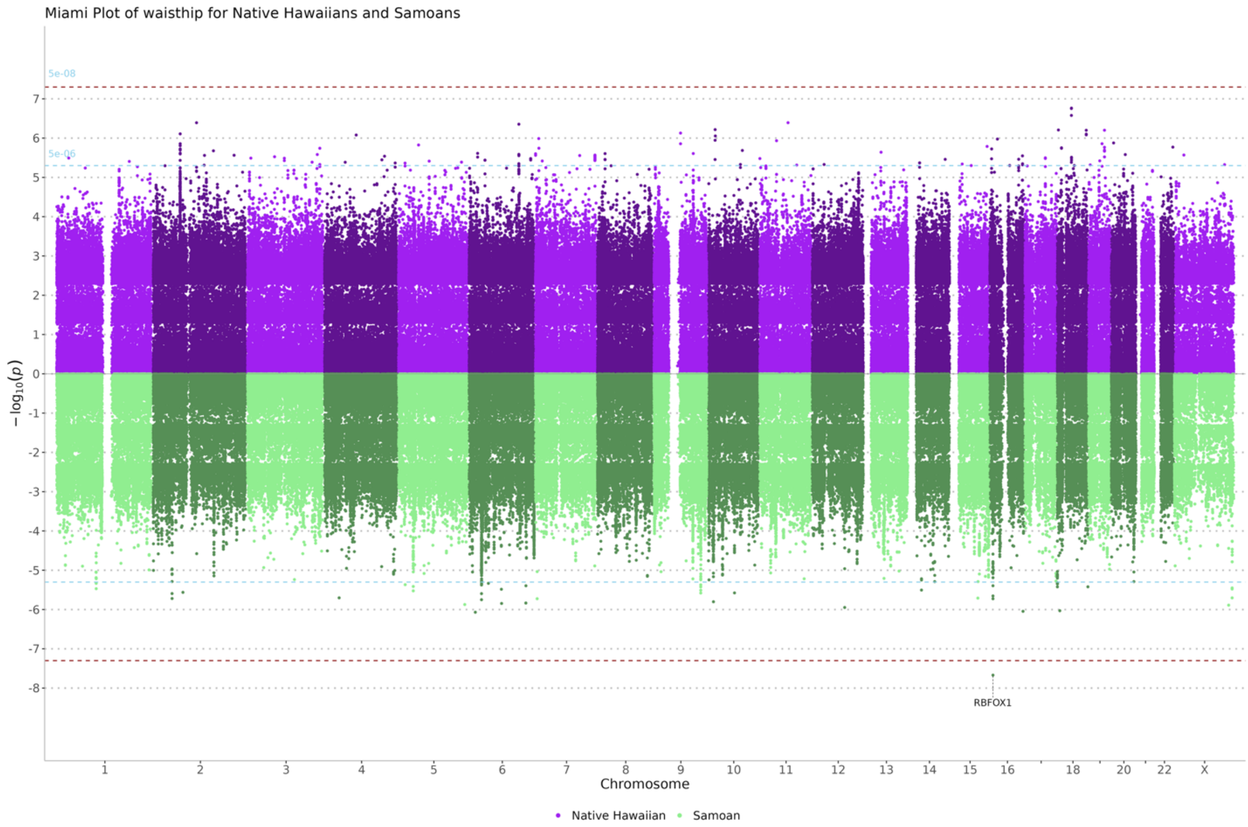


**Supplementary Figure 2. Miami plot of Native Hawaiian and Samoan cohort associations by phenotype.**

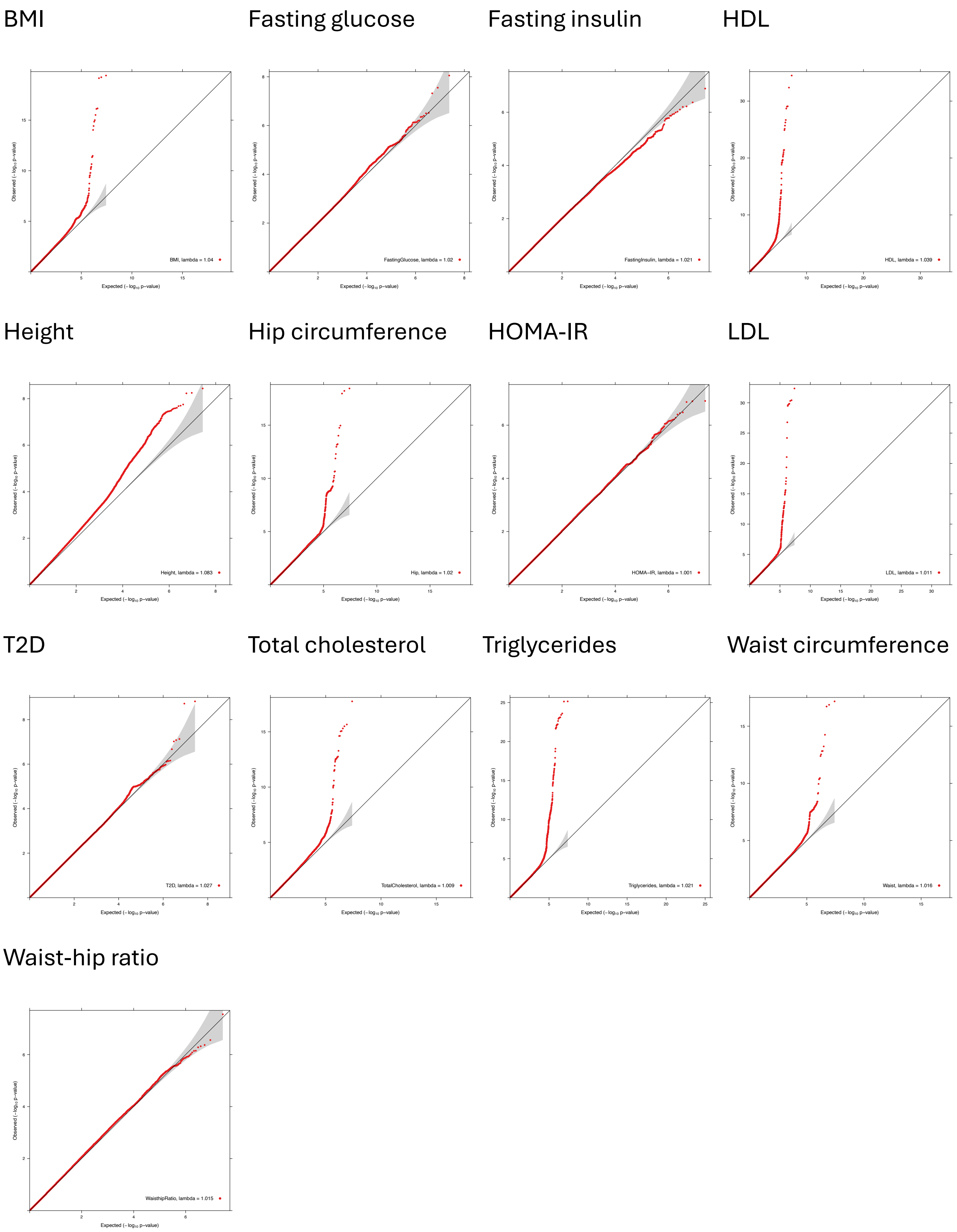


**Supplementary Figure 3. Meta-analysis quantile-quantile plots by trait.**

a. NCOA3 HDL 20:48202897:C:T

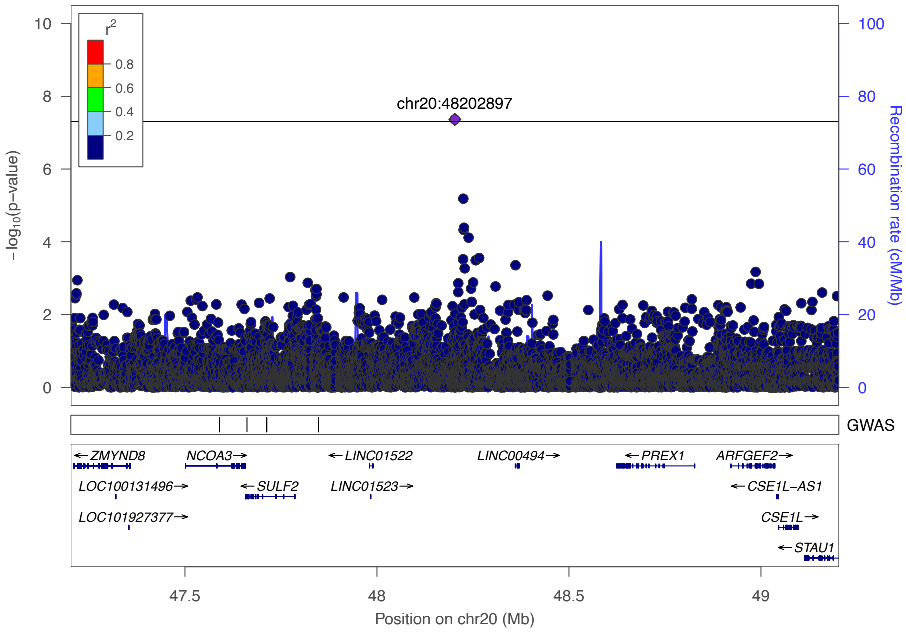


b. APOB total cholesterol 2:21167346:A:G

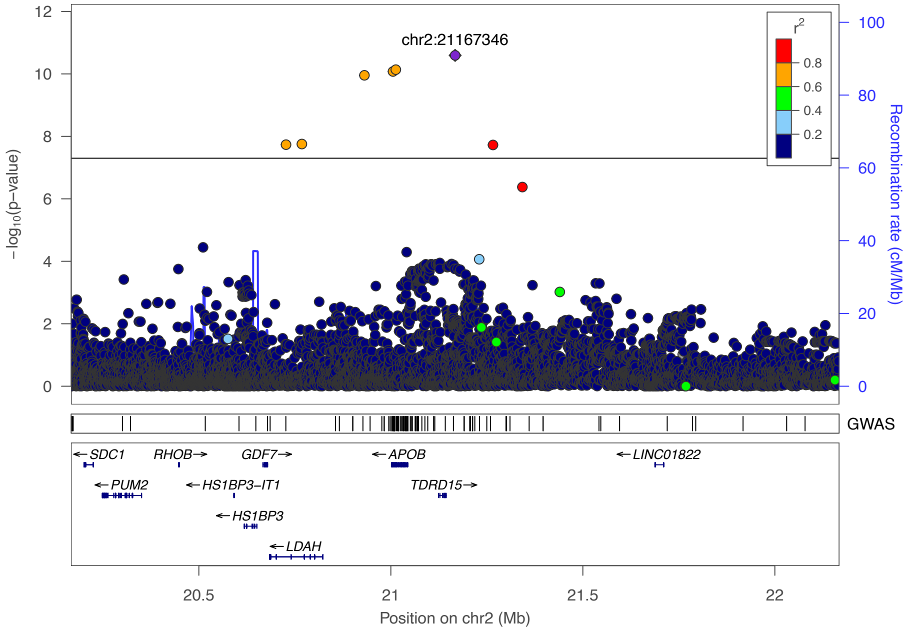

c. APOB LDL 2:21167346:A:G

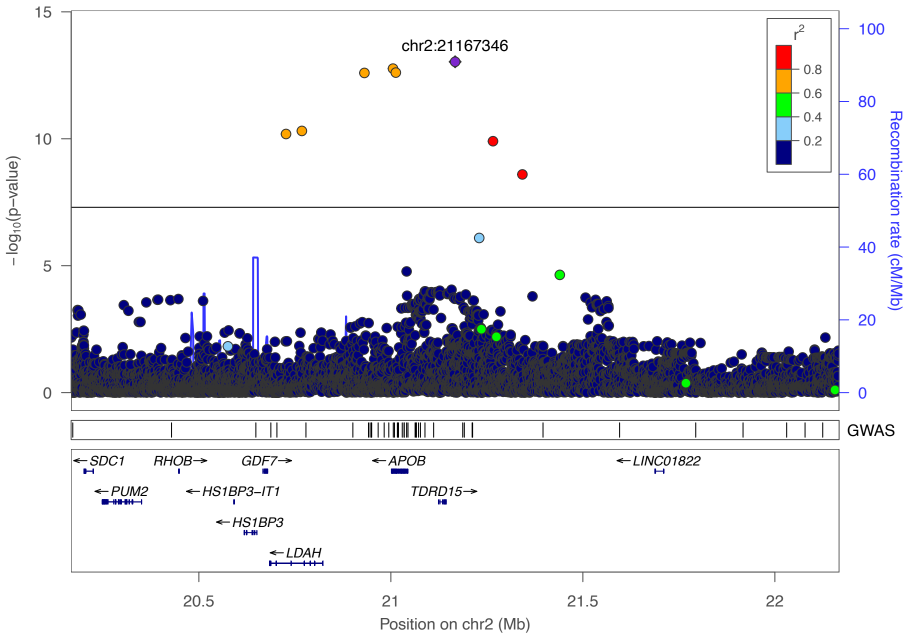


d. HHIP height 4:144412450:A:G

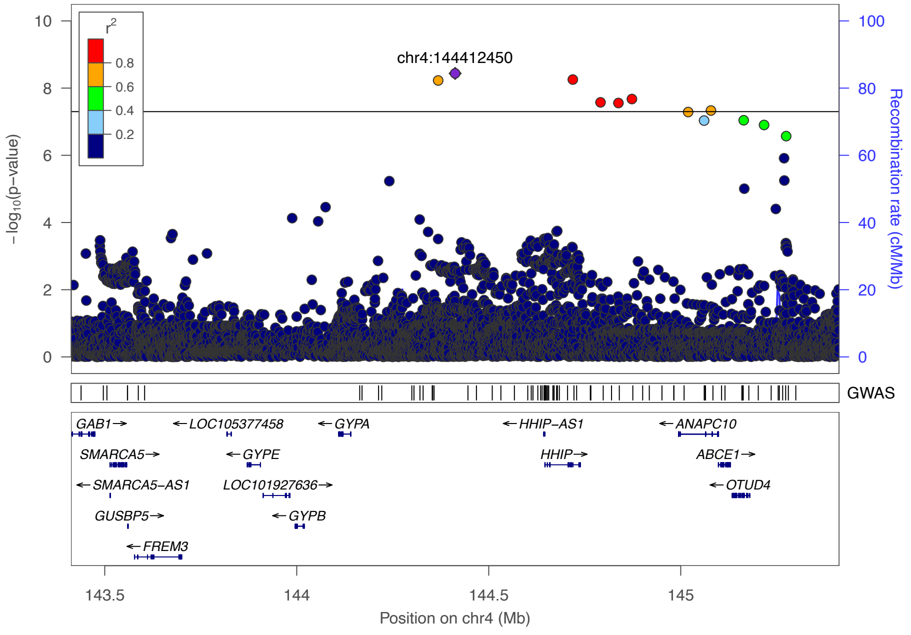

e. LIPG HDL 18:50030280:G:A

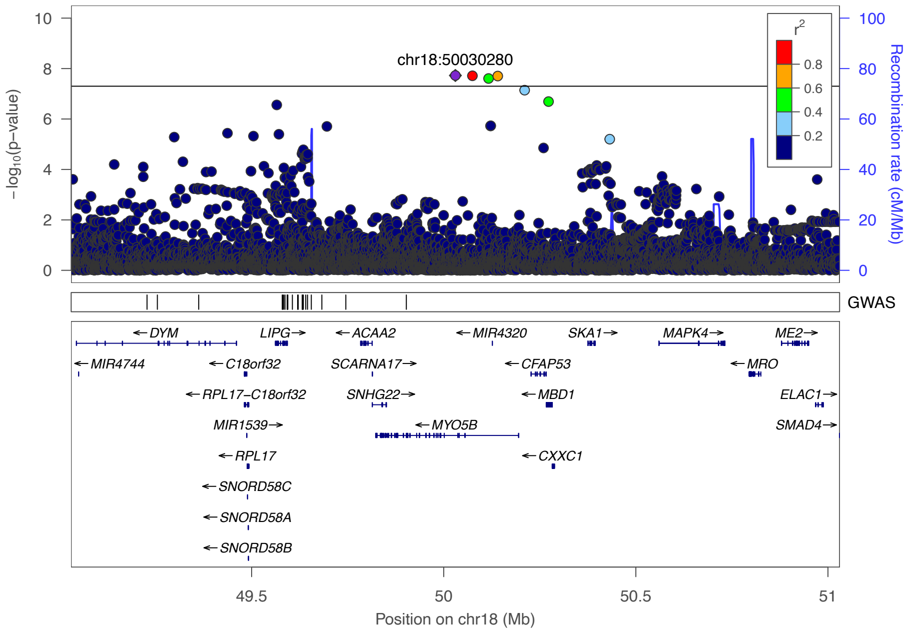


f. ANGPTL8 triglycerides 19:11284678:TC:T

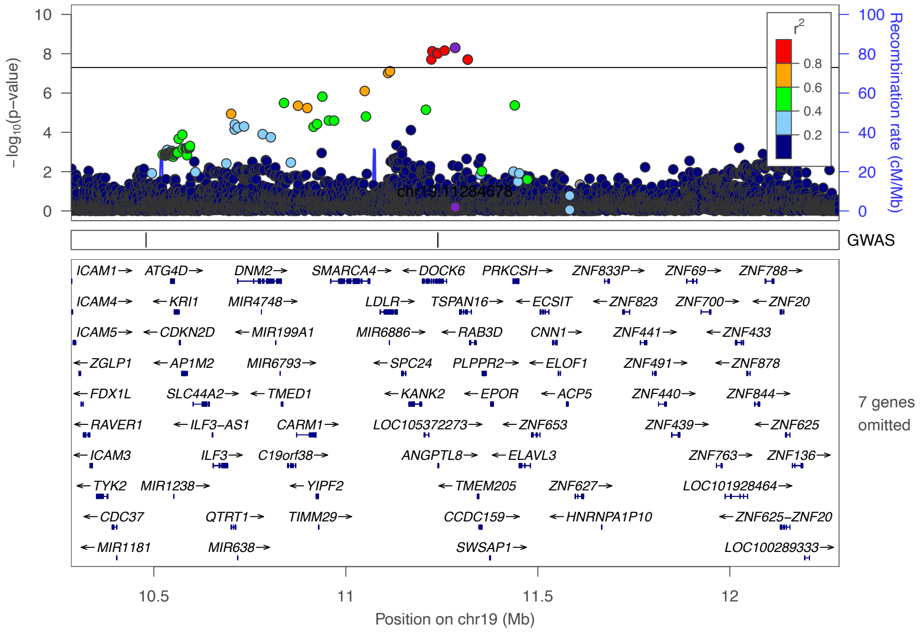

g. APOE LDL 19:44923535:G:A

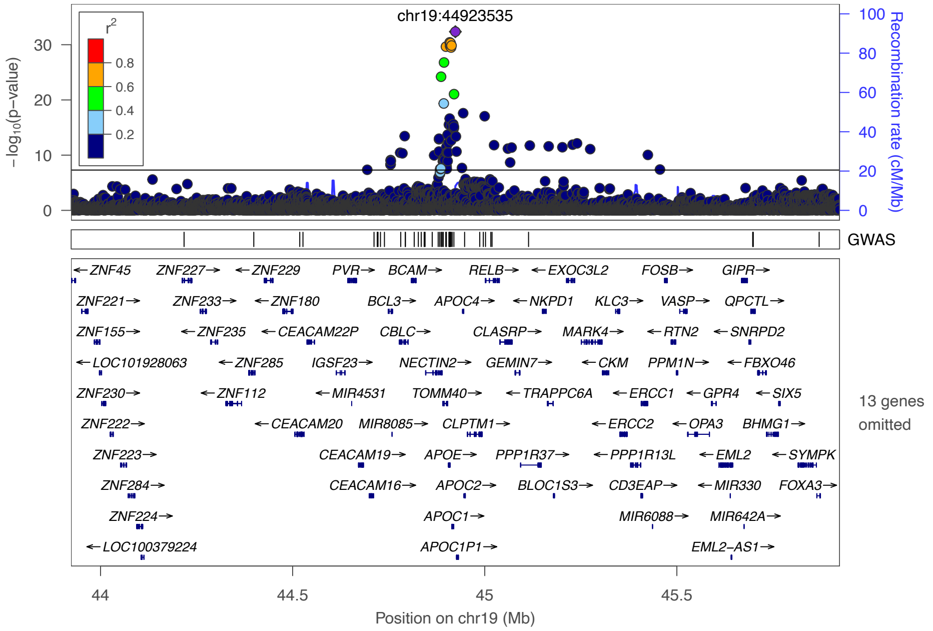


h. APOE total cholesterol 19:44923535:G:A

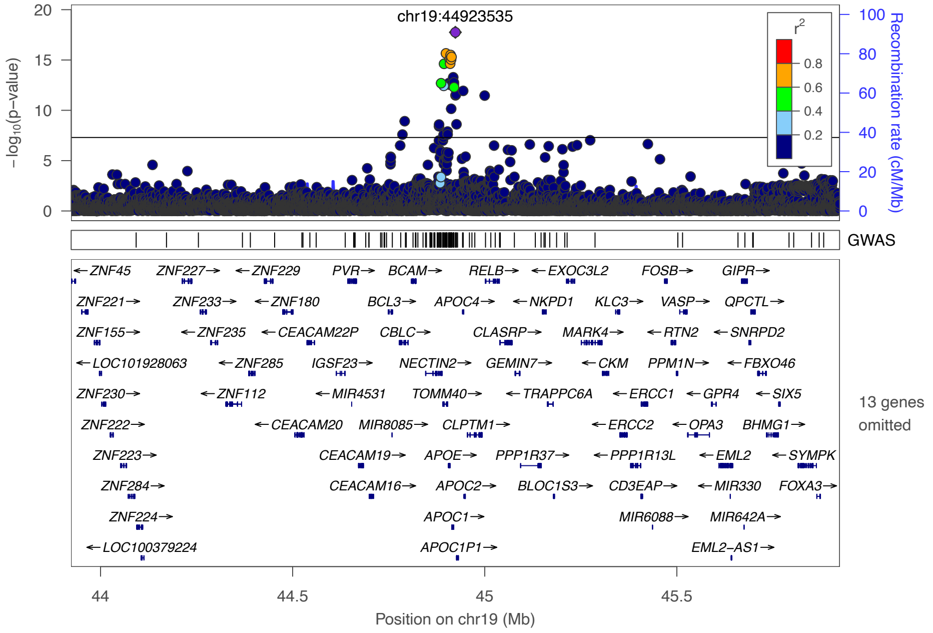

i. VEGFA triglycerides 6:43858616:C:T

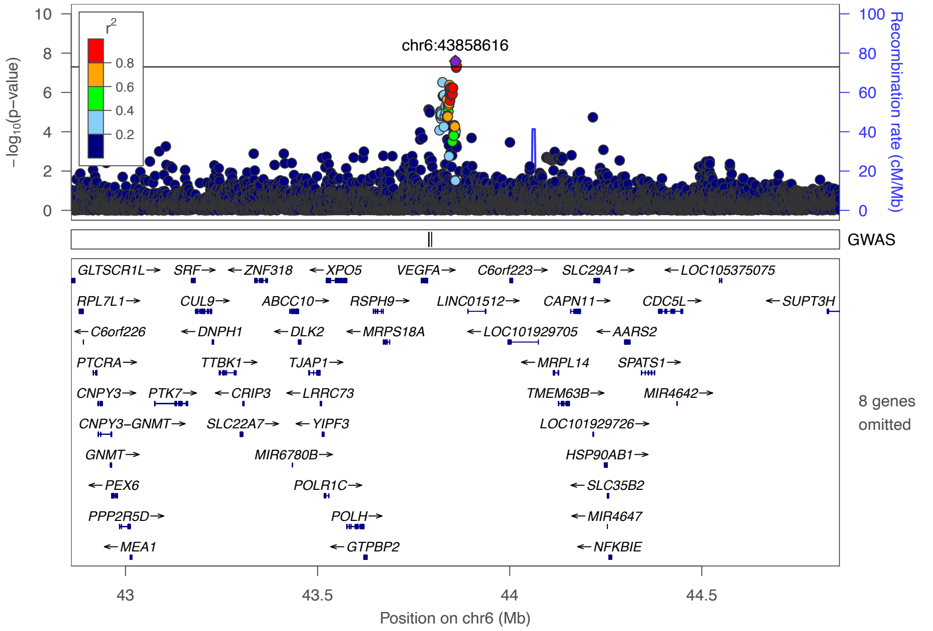


j. APOE HDL 19:44908684:T:C

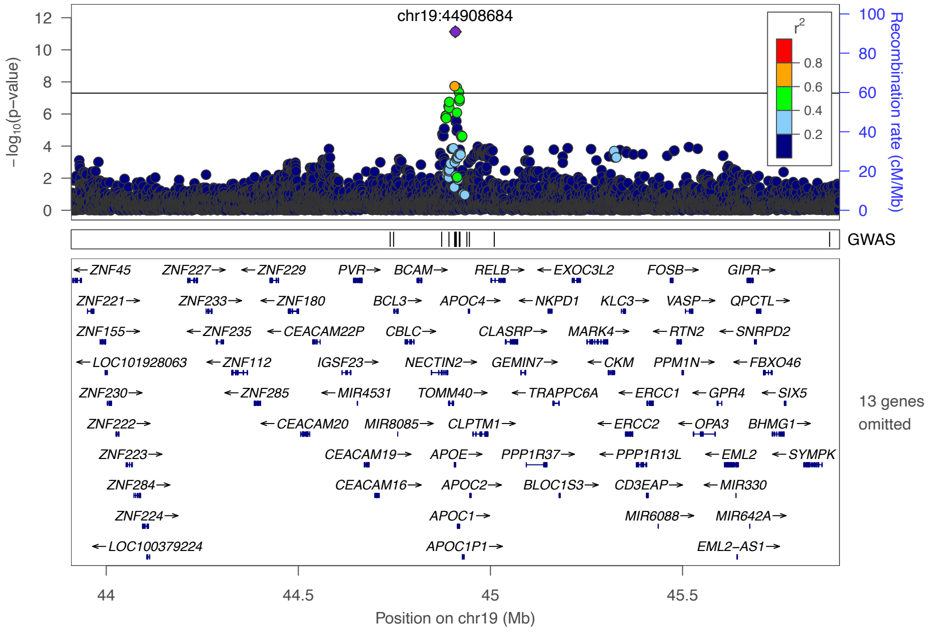

k. PRSS56 height 2:232212354:A:G

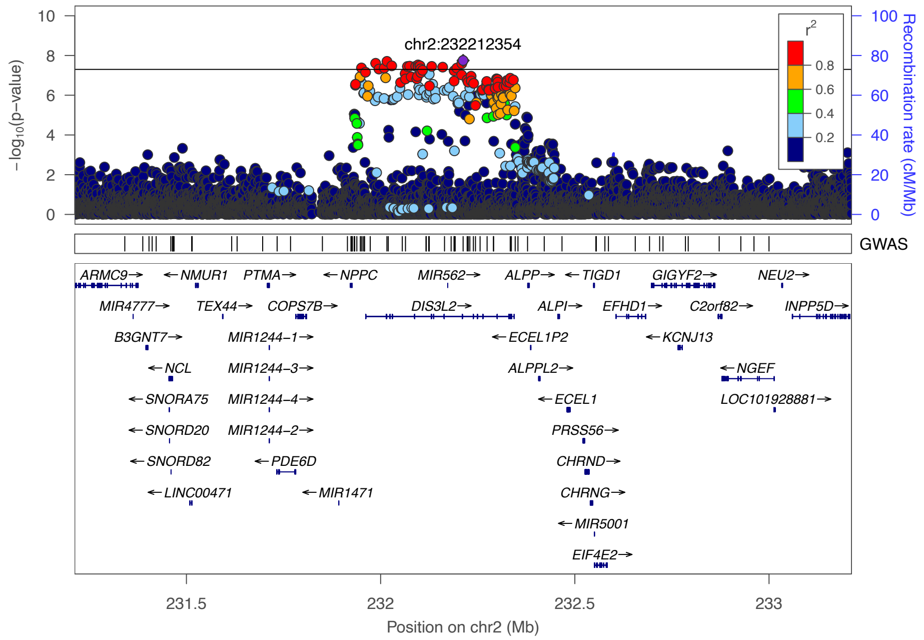


l. APOA5 total cholesterol 11:116786845:C:T

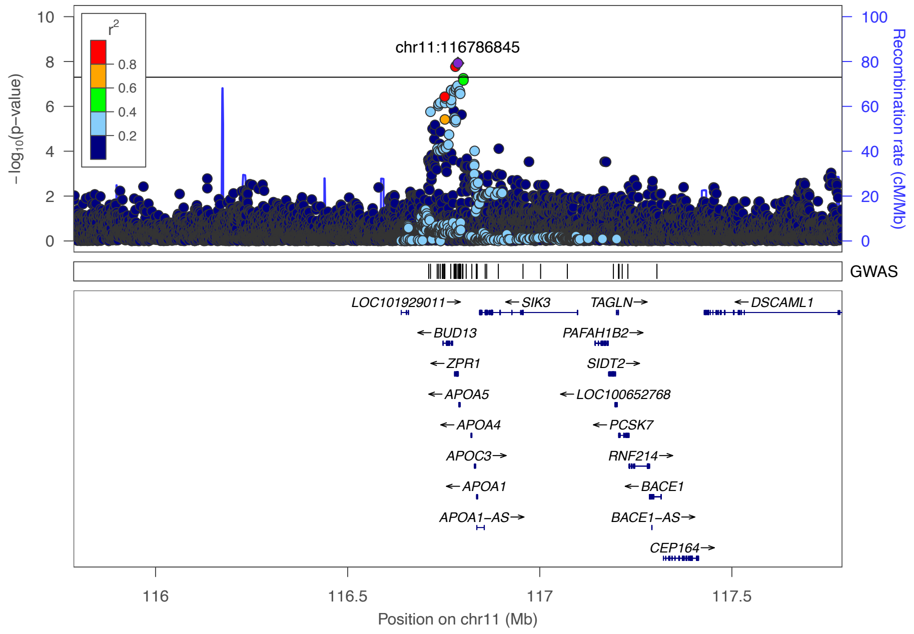

m. APOC4 triglycerides 19:44922203:A:G

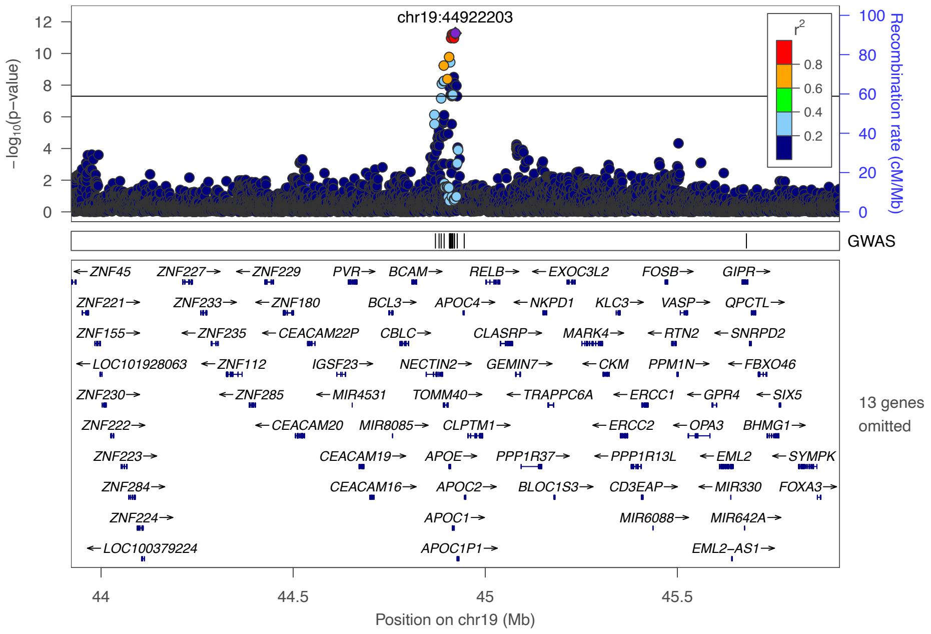


n. GCKR triglycerides 2:27519736:T:C

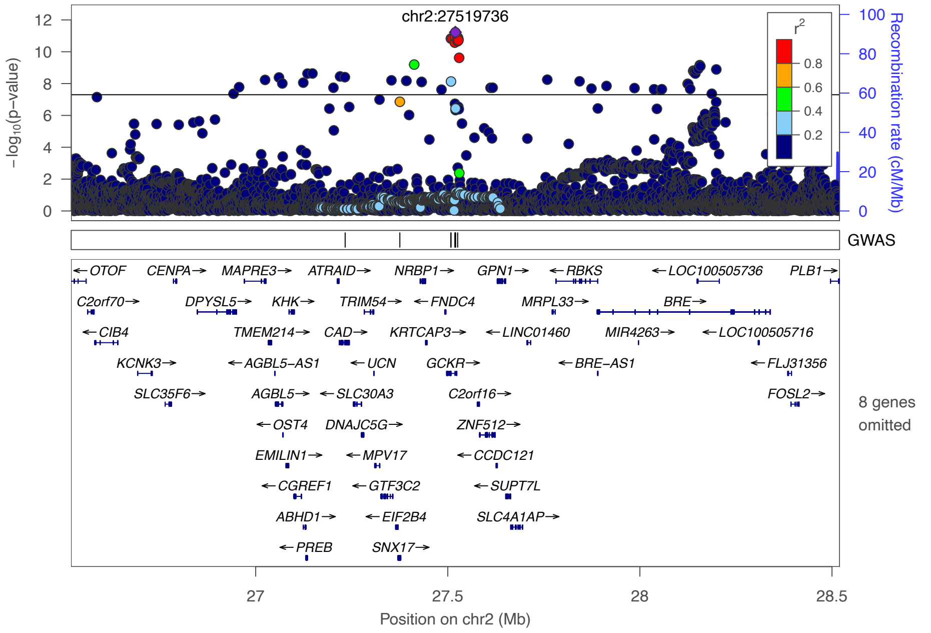

o. APOA5 triglycerides 11:116792991:G:A

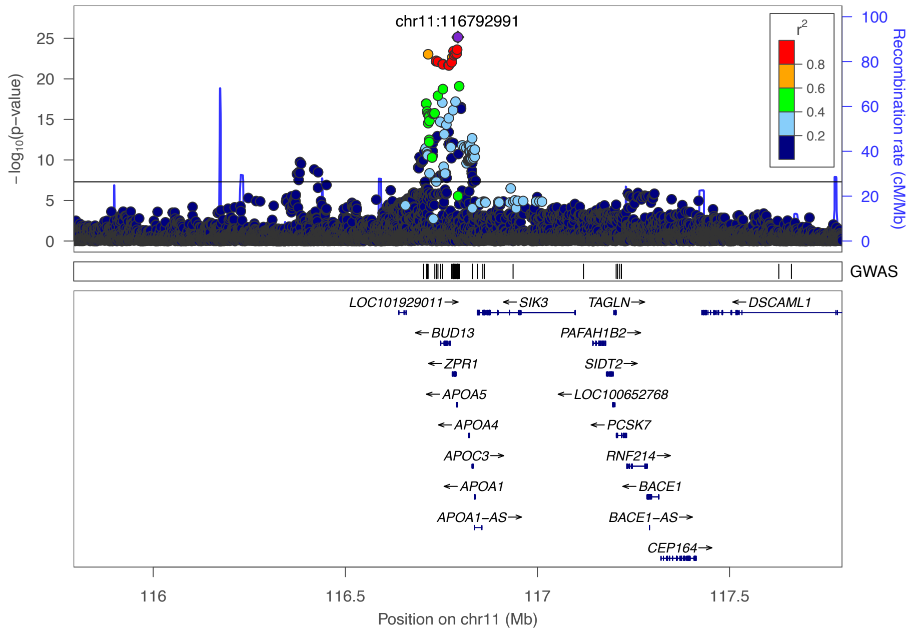


**Supplementary Figure 4. LocusZoom plot for remaining loci with known association based on the GWAS catalog.** The GWAS track shows reported associations in GWAS catalog for the same trait.

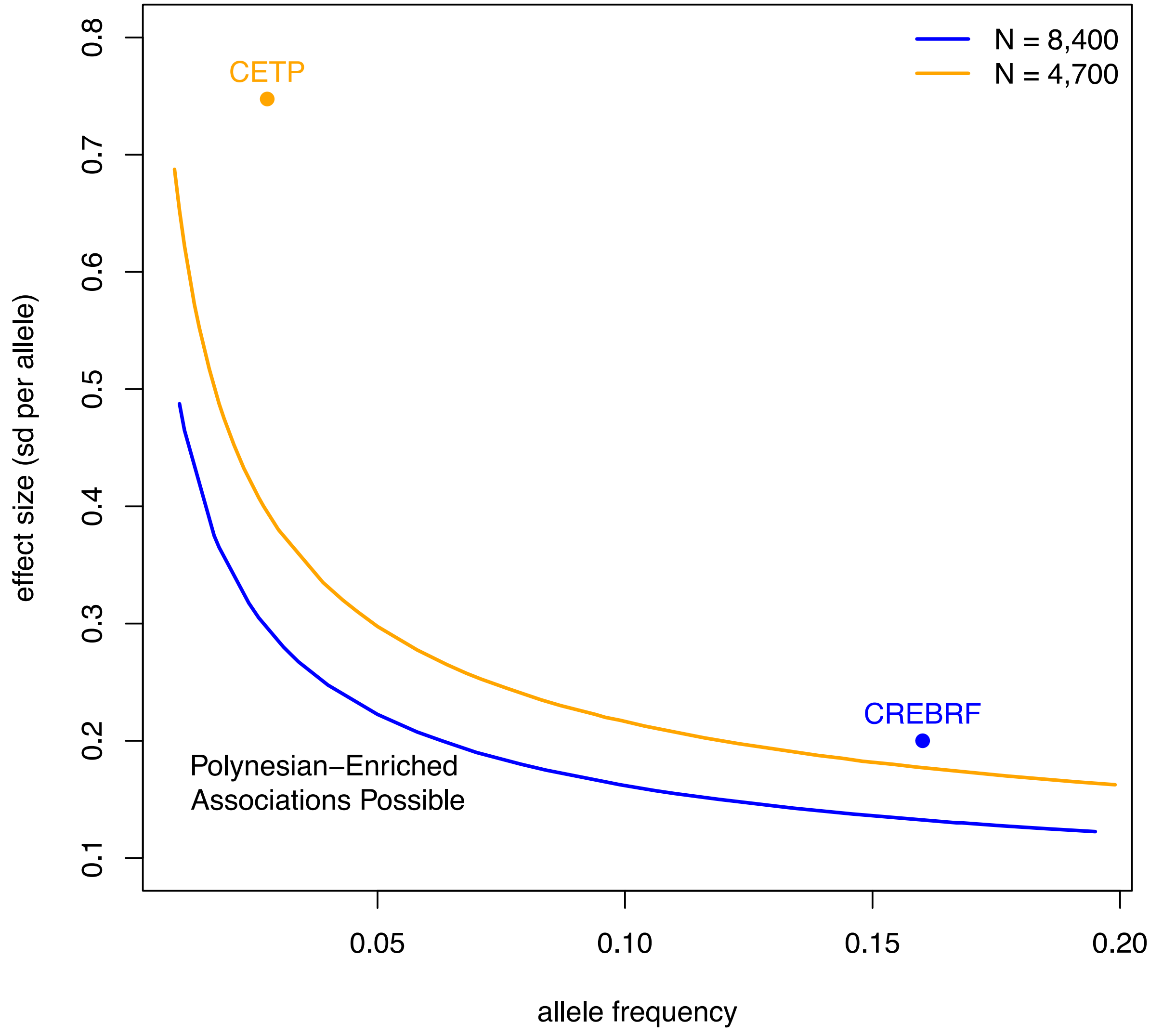


**Supplementary Figure 5. Statistical power of the current study design.** The curves delineates the combination of causal allele frequency and effect sizes (in units of standard deviation, sd, per allele) that could be detected with 80% statistical power in our study assuming a sample size of 8,400 (blue, representing anthropometric traits like BMI) or 4,700 (orange, representing biomarker traits like HDL). For reference, the frequency and effect of the association at the *CREBRF* and *CETP* locus are also shown, colored by their respective trait. Allele frequencies for *CREBRF and CETP* were computed as the weighted average of the Native Hawaiian and Samoan data. It appears that no other common Polynesian-enriched variants exert nearly as large of an effect or segregate at as high of frequency as these two previously found associations.

### Supplemental Tables

| MarkerName | Phenotype | Unconditioned P-value | Conditioned P-value (top hit within 500kb) | When conditioned on CREBRF missense variant |
|---|---|---|---|---|
| **16:7957893:G:C** | Waist-hip ratio | 2.83E-08 | 7.6e-5 | |
| **13:76661594:GTTC:G** | BMI | 4.57E-08 | 2.4e-4 | |
| **2:169265660:G:C** | BMI | 5.68E-09 | 2.4e-4 | |
| **11:98666193:G:GT** | LDL | 6.53E-09 | 2.1e-5 | |
| **5:173045049:C:G** | Fasting glucose | 8.89E-09 | 4.1e-4 | 0.2196 |

**Supplementary Table 1**. **Conditional analysis in newly confirmed loci**. Two separate conditional analyses were performed for the loci with potentially novel associations. First, in order to detect potential secondary signals, we conditioned on the lead variant at each locus and report the smallest p-value within 500kb. No additional signals were detected from this analysis. Second, in order to determine if the signal at the CREBRF locus is in fact reflecting the known missense variant, we conditioned on rs373863828 (chr5:173108771:C:G). We find that the signal at the locus decreases significantly and conclude that the original signal is due to the CREBRF missense variant.

| MarkerName | Phenotype | Unconditioned P-value | Conditioned P-value (top hit within 500kb) |
|---|---|---|---|
| **20:48202897:C:T** | HDL | 4.34E-08 | 0.1414 |
| **2:21167346:A:G** | Total cholesterol | 2.55E-11 | 0.07814 |
| **2:21167346:A:G** | LDL | 9.28E-14 | 0.0946 |
| **4:144412450:A:G** | Height | 3.65E-09 | 0.919 |
| **18:50030280:G:A** | HDL | 1.88E-08 | 0.7099 |
| **16:56971035:C:T** | HDL | 3.25E-35 | 0.001463 |
| **19:11284678:TC:T** | Triglycerides | 4.99E-09 | 0.0005196 |
| **19:44923535:G:A** | LDL | 4.41E-33 | 2.62E-09 |
| **19:44923535:G:A** | Total cholesterol | 1.77E-18 | 5.53E-06 |
| **5:173045049:C:G** | BMI | 3.91E-20 | 0.4646 |
| **6:43858616:C:T** | Triglycerides | 2.57E-08 | 0.9401 |
| **19:44908684:T:C** | HDL | 7.52E-12 | 0.9739 |
| **2:232212354:A:G** | Height | 1.79E-08 | 0.1292 |
| **11:116786845:C:T** | Total cholesterol | 1.19E-08 | 0.619 |
| **19:44922203:A:G** | Triglycerides | 5.27E-12 | 0.000196 |
| **2:27519736:T:C** | Triglycerides | 6.01E-12 | 0.4196 |
| **11:116792991:G:A** | Triglycerides | 7.08E-26 | 0.03322 |
| **5:173045049:C:G** | Waist circumference | 6.82E-18 | 7.4e-5 |
| **5:173045049:C:G** | Hip circumference | 3.56E-19 | 1.0e-5 |

**Supplementary Table 2. Conditional analysis at loci with previously known associations.** Conditional analyses were conducted using the dosage of the previously reported variant in the GWAS catalog. If multiple nearby hits were found, the variant with the strongest association in our meta-analysis was used. Across all loci, association signals decreased drastically after conditional analysis leading us to conclude that the initial signals reflect previously reported findings.

| Phenotype | Plausible Gene | chr | pos | SNP type AoU | AoU ancestry | AoU beta | AoU pval | AoU N | AoU AF | Effect (meta) | pval (meta) | NH p | NH effect | Samoan p | Samoan effect | h2 | power |
|---|---|---|---|---|---|---|---|---|---|---|---|---|---|---|---|---|---|
| **BMI** | **MYCBP2** | chr13 | 76661594 | top SNP | AFR | -0.0119 | 0.193 | 49885 | 0.1266 | 1.03 | 4.6E-08 | 8.0E-08 | -1.14 | 1.2E-01 | -0.62 | 0.234 | 100.00% |
| | | | | | AMR | -0.0142 | 0.637 | 38834 | 0.0148 | | | | | | | 0.0308 | 100.00% |
| | | | | | EUR | 0.174 | 0.043 | 111482 | 0.0006 | | | | | | | 0.00128 | 100.00% |
| | | | | | SAS | 0.555 | 0.127 | 2892 | 0.0012 | | | | | | | 0.00256 | 55.75% |
| **BMI** | **BBS5** | chr2 | 169265660 | top SNP | AFR | 0.0137 | 0.331 | 49885 | 0.0485 | 0.460 | 5.7E-09 | 3.9E-08 | -0.44 | 3.1E-02 | -0.81 | 0.0195 | 100.00% |
| | | | | | AMR | -0.00176 | 0.930 | 38834 | 0.0324 | | | | | | | 0.0132 | 100.00% |
| | | | | | EUR | -0.00546 | 0.604 | 111482 | 0.0411 | | | | | | | 0.0167 | 100.00% |
| | | | | | SAS | -0.0164 | 0.800 | 2892 | 0.0399 | | | | | | | 0.0162 | 100.00% |
| **fasting glucose** | **CREBRF** | chr5 | 173045049 | top SNP | AFR | 0.00239 | 0.982 | 24763 | 0.0017 | -0.0964 | 8.9E-09 | 2.8E-04 | -0.07 | 3.2E-07 | -0.17 | 1.83E-05 | 2.91% |
| | | | | | AMR | -0.00655 | 0.704 | 21811 | 0.0724 | | | | | | | 0.000716 | 91.55% |
| | | | | | EAS | 0.0374 | 0.496 | 2271 | 0.0669 | | | | | | | 0.000666 | 8.92% |
| | | | | | EUR | -0.0103 | 0.936 | 73568 | 0.0004 | | | | | | | 3.98E-06 | 2.19% |
| | | | | | SAS | -0.206 | 0.306 | 1355 | 0.0085 | | | | | | | 8.97E-05 | 1.47% |
| | | | | | AFR | 0.0780 | 0.871 | 24763 | 0.0001 | | | | | | | 8.61E-07 | 1.08% |
| **LDL** | **CNTN5** | chr11 | 98666193 | top SNP | AFR | 0.280 | 0.572 | 12558 | 0.0002 | 15.5 | 6.5E-09 | 1.1E-4 | 20.27 | 8.6E-06 | 13.86 | 5.87E-05 | 4.33% |
| | | | | | AMR | 0.774 | 0.262 | 9551 | 0.0001 | | | | | | | 3.86E-05 | 2.52% |
| | | | | | EUR | -1.31 | 0.177 | 40699 | 0.0000 | | | | | | | 4.53E-06 | 1.72% |
| **Waist-hip ratio** | **RBFOX1** | chr16 | 7957893 | top SNP | AFR | 0.138 | 0.691 | 46963 | 0.0001 | 0.538 | 2.8E-08 | 3.9E-01 | -0.24 | 2.1E-08 | -0.58 | 4.92E-05 | 14.56% |
| | | | | | AMR | 0.0736 | 0.878 | 34995 | 0.0001 | | | | | | | 3.30E-05 | 6.69% |
| | | | | | EAS | -0.152 | 0.245 | 4762 | 0.0046 | | | | | | | 0.00266 | 83.70% |
| | | | | | SAS | -0.000507 | 0.997 | 2698 | 0.0097 | | | | | | | 0.00553 | 90.07% |

**Supplementary Table 3. Power to replicate potentially novel variant-trait association in All-of-Us (AoU).** We calculated power to replicate a finding in AoU assuming effect sizes in units of standard deviation from our meta-analysis, but using frequency and sample size of the AoU cohort for replication. The critical value threshold used in power calculation was 6.6349, which was calculated for P = 0.01. For fasting glucose and LDL the units of the associated results were converted into units of standard deviation. Most of the phenotypes from the novel loci-phenotype pairs are available in AoU, with the exception of waist-to-hip ratio; for waist-hip ratio, we use mean waist circumference, a related phenotype for replication. As previously reported, a CREBRF variant (rs12513649) is found more commonly across global populations in AoU, while the causal missense variant (rs373863828) is extremely rare. Rs12513649 is in LD with rs373863828 among Polynesians and thus was the top associated variant in our study. However, despite being powered to detect associations with rs12513649 in AMR and EAS populations, it is not replicated in AoU likely because the variant is not in LD with the causal variants outside of Polynesia. Rs373863828 was not tested by AoU due to its extreme rarity. Lastly, for the AFR subcohort in AoU, associations with rs12513649 are testing the T alternative allele rather than the G allele, which is not available. AFR – African ancestry , AMR – Admixed American, EAS – East Asian, SAS – South Asian, EUR – European.

| Phenotype | Units | Covariates | Transformation | Excluded |
|---|---|---|---|---|
| **BMI** | Ratio of weight (kg) to height (cm) squared | Sex specific: age | INT | Ind. < 18 years old, ind. > 6 s.d. |
| **Fasting glucose** | mmol/L | Age, sex, age*sex, smoking status, BMI | | Ind. pregnant, fasting glucose > 7 mmol/L, had T2D, or not fasting at measurement |
| **Fasting insulin** | pmol/L | Age, sex, age*sex, smoking status, BMI | | Ind. pregnant, fasting glucose > 7 mmol/L, had T2D, or not fasting at measurement |
| **HDL** | mg/dL | Age, sex | | Ind. pregnant at blood draw or non-fasting |
| **Height at enrollment** | cm | Sex specific: age | INT | Ind. < 18 years old, ind. > 6 s.d. |
| **Hip circumference** | cm | Sex specific: age | INT | Ind. < 18 years old, ind. > 6 s.d. |
| **HOMA-IR** | Insulin*Glucose/22.5 | Age, sex, age*sex, smoking status, BMI | | Ind. pregnant, fasting glucose > 7 mmol/L, had T2D, or not fasting at measurement |
| **LDL** | mg/dL | Age, sex | Friedewald equation | Ind. pregnant, fasted < 8 hours before lipid blood draw, TG value > 400 mg/dL |
| **Total Cholesterol** | mg/dL | Age, sex | | Ind. pregnant, fasted < 8 hours before lipid blood draw |
| **Triglycerides** | mg/dL | Age, sex | Natural log | Ind. pregnant, fasted < 8 hours before lipid blood draw, TG value > 3000 mg/dL |
| **Waist circumference** | cm | Sex specific: age | INT | Ind. < 18 years old, ind. > 6 s.d. |
| **Waist-to-hip ratio** | Waist/Hip | Sex specific: age, BMI | INT | Ind. < 18 years old, ind. > 6 s.d. |

**Supplementary Table 4. Exclusion criteria, units, and covariates for phenotype transformation are listed for tested quantitative traits.** The number of PCs included in the linear mixed model, in addition to a GRM, is 20 for the Native Hawaiian cohort and 3 for the Samoan cohort. INT – inverse normal transformed.
