## Supplemental Text for "Meta-analysis of over 8,000 individuals from Hawai‘i and Samoa for genetic associations to cardiometabolic phenotypes"

### Supplementary Text

**Artificial intelligence-based literature search to nominate plausible associated gene at a genomic locus.**
For every associated region, we used the following prompt (in `Courrier New`) to query ChatGPT 4o mini to nominate plausible gene in the genomic locus responsible for the observed genetic association. We also record the output (in blue) from ChatGPT. All queries shared the same initial prompt:

```
You are an expert in biology and genetics. Your task is to
identify likely causal genes within a locus for a given GWAS
phenotype based on literature evidence.

From the list, provide the likely causal gene (matching one of
the genes), confidence (0: very unsure to 1: very confident),
and a brief reason (50 words or less) for your choice.

Identify the causal gene.
```

The standard query was then followed by locus-specific information of the associated trait and list of potential genes around the locus.

**Prompt and Output for Potentially Novel Loci:**

```
GWAS: waist-to-hip ratio

Genes in locus: {RBFOX1}, {TMEM114}, {TMEM186}, {METTL22},
{USP7}, {ABAT}, {PMM2}, {CARHSP1}
```

**Likely causal gene**: **RBFOX1**
**Confidence**: 0.8
**Reason**: RBFOX1 is known to regulate RNA splicing and has been implicated in metabolic processes, including fat distribution. Its role in adipocyte differentiation and fat metabolism makes it a plausible candidate for waist-to-hip ratio phenotypes, as supported by functional studies in metabolic and obesity-related traits.

```
GWAS: BMI

Genes in locus: {LMO7}, {KCTD12}, {MYCBP2}, {SCEL}, {LMO7DN},
{LMO7DN-IT1}, {BTF3P11}, {SCEL-AS1}, {ACOD1}, {LOC100129307},
{CLN5}, {FBXL3}, {MYCBP2-AS1}
```

**Likely causal gene**: **MYCBP2**

**Confidence**: **0.8**

**Reason**: MYCBP2 (MYC binding protein 2) is implicated in processes related to cellular growth and metabolism, which are linked to BMI regulation. There is evidence suggesting MYCBP2 influences pathways that control adipogenesis and energy balance, making it a strong candidate for BMI-associated phenotypes.

GWAS: BMI

Genes in locus: {CERS6}, {SPC25}, {DHRS9}, {BBS5}, {KLHL23}, {MYO3B}, {MIR4774}, {NOSTRIN}, {LRP2}, {KLHL41}, {SSB}, {CERS6-AS1}, {FASTKD1}, {METTL5}, {G6PC2}, {PPIG}, {SNORD3K}, {ABCB11}, {CCDC173}, {UBR3}, {PHOSPHO2-KLHL23}, {PHOSPHO2}

**Causal gene**: **BBS5**
**Confidence**: **0.85**
**Reason**: BBS5 is associated with Bardet-Biedl Syndrome, a disorder that includes obesity as a key feature. The gene is involved in cilia function, which regulates energy balance and fat storage. Several studies have linked BBS5 mutations to BMI-related traits.

GWAS: LDL
Genes in locus: {CNTN5}

**Likely Causal Gene:** CNTN5
**Confidence:** 0.7
**Reason:** CNTN5 (Contactin 5) is a member of the contactin family, involved in cell adhesion and signaling, which could affect lipid metabolism. While direct evidence for LDL is limited, its involvement in neuronal and endothelial signaling suggests a plausible link to lipid regulation, warranting moderate confidence.

GWAS: waist circumference

Genes in locus: {STK10}, {SH3PXD2B}, {NEURL1B}, {ERGIC1}, {BNIP1}, {STC2}, {LOC285593}, {CPEB4}, {EFCAB9}, {MIR5003}, {LOC100268168}, {NKX2-5}, {BOD1}, {C5orf47}, {UBTD2}, {LOC101928093}, {CREBRF}, {MIR8056}, {LINC01484}, {LOC100288254}, {DUSP1}, {ATP6V0E1}, {LINC01485}, {RPL26L1}, {SNORA74B}

**Likely causal gene: CREBRF**
**Confidence: 0.85**
**Reason:** CREBRF has been implicated in obesity-related traits in GWAS, including waist circumference, through its association with adiposity and metabolic traits. It has a known role in regulating energy balance and is functionally relevant to body composition.

GWAS: hip circumference

Genes in locus: {STK10}, {SH3PXD2B}, {NEURL1B}, {ERGIC1}, {BNIP1}, {STC2}, {LOC285593}, {CPEB4}, {EFCAB9}, {MIR5003}, {LOC100268168}, {NKX2-5}, {BOD1}, {C5orf47}, {UBTD2}, {LOC101928093}, {CREBRF}, {MIR8056}, {LINC01484}, {LOC100288254}, {DUSP1}, {ATP6V0E1}, {LINC01485}, {RPL26L1}, {SNORA74B}

**Likely Causal Gene:** **CREBRF**
**Confidence:** 0.85
**Reason:** CREBRF has been implicated in metabolic traits, including body size and adiposity, which are closely related to hip circumference. Genetic variations in CREBRF have shown associations with obesity and fat distribution, suggesting a plausible link to hip circumference.

GWAS: fasting glucose

Genes in locus: {STK10}, {SH3PXD2B}, {NEURL1B}, {ERGIC1}, {BNIP1}, {STC2}, {LOC285593}, {CPEB4}, {EFCAB9}, {MIR5003}, {LOC100268168}, {NKX2-5}, {BOD1}, {C5orf47}, {UBTD2}, {LOC101928093}, {CREBRF}, {MIR8056}, {LINC01484}, {LOC100288254}, {DUSP1}, {ATP6V0E1}, {LINC01485}, {RPL26L1}, {SNORA74B}

**Likely causal gene: CREBRF**
**Confidence: 0.8**
**Reason:** CREBRF has been associated with glucose metabolism and insulin resistance in several studies, particularly in relation to fasting glucose levels. It plays a role in the regulation of metabolic processes, making it a strong candidate for the fasting glucose phenotype in GWAS.

**Prompt and Output for Previously Known Loci:**

GWAS: HDL

Genes in locus: {ZMYND8}, {NCOA3}, {LINC01522}, {LINC00494}, {PREX1}, {ARFGEF2}, {LOC100131496}, {SULF2}, {LINC01523}, {CSE1L-AS1}, {LOC101927377}, {CSE1L}, {STAU1}

**Likely Causal Gene: NCOA3**
**Confidence: 0.85**
**Reason:** NCOA3 (Nuclear Receptor Coactivator 3) is known to play a role in lipid metabolism and is associated with regulation of HDL levels. It functions as a coactivator for nuclear receptors that influence lipid-related gene expression, making it a strong candidate for the GWAS phenotype.

GWAS: total cholesterol

Genes in locus: {SDC1}, {RHOB}, {GDF7}, {APOB}, {LINC01822}, {PUM2}, {HS1BP3-IT1}, {TDRD15}, {HS1BP3}, {LDAH}

**Causal Gene: APOB**

**Confidence: 0.9**

**Reason**: APOB (Apolipoprotein B) is directly involved in lipid metabolism and is a known major gene affecting cholesterol levels. Variants in APOB are well-documented to influence total cholesterol and lipid-related phenotypes, making it the most likely causal gene in this locus for the given trait.

GWAS: LDL

Genes in locus: {SDC1}, {RHOB}, {GDF7}, {APOB}, {LINC01822}, {PUM2}, {HS1BP3-IT1}, {TDRD15}, {HS1BP3}, {LDAH}

**Causal Gene**: **APOB**
**Confidence**: 0.9
**Reason**: APOB is a key gene in lipid metabolism and a well-established player in LDL cholesterol regulation. Variants in APOB have been directly linked to changes in LDL levels, making it the most likely causal gene in this locus for the GWAS phenotype.

GWAS: height

Genes in locus: {GAB1}, {LOC105377458}, {GYPA}, {HHIP-AS1}, {ANAPC10}, {SMARCA5}, {GYPE}, {HHIP}, {ABCE1}, {SMARCA5-AS1}, {LOC101927636}, {GUSBP5}, {GYPB}, {OTUD4}, {FREM3}

**Likely Causal Gene:** HHIP
**Confidence:** 0.85
**Reason:** HHIP (Hedgehog Interacting Protein) is known to regulate growth and development through the hedgehog signaling pathway, which is implicated in height. Several GWAS studies have pointed to HHIP as a candidate gene for height-related traits due to its role in skeletal growth.

GWAS: HDL

Genes in locus: {DYM}, {LIPG}, {ACAA2}, {MIR4320}, {SKA1}, {MAPK4}, {ME2}, {MIR4744}, {C18orf32}, {SCARNA17}, {CFAP53}, {MRO}, {RPL17-C18or32}, {SNHG22}, {MBD1}, {ELAC1}, {MIR1539}, {MYO5B}, {SMAD4}, {RPL17}, {CXXC1}, {SNORD58C}, {SNORD58A}, {SNORD58B}

**Likely Causal Gene:** **LIPG**
**Confidence:** 0.9
**Reason:** LIPG (endothelial lipase) is a well-established gene involved in HDL metabolism and regulation. It has been frequently implicated in lipid traits, including HDL levels, in multiple GWAS studies. Other genes in the list are less directly linked to HDL metabolism.

GWAS: HDL

Genes in locus: {LOC283856}, {AMFR}, {MT4}, {NUP93}, {CETP}, {FAM192A}, {CCL17}, {ADGRG1}, {CNGB1}, {GNAO1}, {BBS2}, {MT1G}, {SLC12A3}, {CPNE2}, {CCL22}, {ADGRG3}, {DKFZP434H168}, {MT3}, {MIR138-2}, {RSPRY1}, {CIAPIN1}, {DRC7}, {MIR3935}, {MT2A}, {MIR6863}, {ARL2BP}, {COQ9}, {KATNB1}, {NUDT21}, {MT1E}, {HERPUD1}, {PLLP}, {POLR2C}, {KIFC3}, {OGFOD1}, {MT1F}, {NLRC5}, {CX3CL1}, {MIR6772}, {MT1L}, {DOK4}, {LOC388282}, {MT1M}, {CCDC102A}, {MT1JP}, {ADGRG5}, {MT1A}

**Causal Gene:** **CETP**
**Confidence:** 0.9
**Reason:** CETP (cholesteryl ester transfer protein) is a well-established gene associated with HDL cholesterol metabolism. Multiple GWAS studies have demonstrated its role in regulating HDL levels and lipid homeostasis. Given its direct involvement in lipid transport, it is the most likely causal gene in this locus.

GWAS: triglycerides

Genes in locus: {ICAM1}, {ATG4D}, {DNM2}, {SMARCA4}, {DOCK6}, {PRKCSH}, {ZNF833P}, {ZNF69}, {ZNF788}, {ICAM4}, {KRI1}, {MIR4748}, {LDLR}, {TSPAN16}, {ECSIT}, {ZNF823}, {ZNF700}, {ZNF20}, {ICAM5}, {CDKN2D}, {MIR199A1}, {MIR6886}, {RAB3D}, {CNN1}, {ZNF441}, {ZNF433}, {ZGLP1}, {AP1M2}, {MIR6793}, {SPC24}, {PLPPR2}, {ELOF1}, {ZNF491}, {ZNF878}, {FDX1L}, {SLC44A2}, {TMED1}, {KANK2}, {EPOR}, {ACP5}, {ZNF440}, {ZNF844}, {RAVER1}, {ILF3-AS1}, {CARM1}, {LOC105372273}, {ZNF653}, {ZNF439}, {ZNF625}, {ICAM3}, {ILF3}, {C19orf38}, {ANGPTL8}, {ELAVL3}, {ZNF763}, {ZNF136}, {TYK2}, {MIR1238}, {YIPF2}, {TMEM205}, {ZNF627}, {LOC101928464}, {CDC37}, {QTRT1}, {TIMM29},

{CCDC159}, {HNRNPA1P10}, {ZNF625-ZNF20}, {MIR1181}, {MIR638}, {SWSAP1}, {LOC100289333}

**Likely causal gene:** **ANGPTL8**
**Confidence:** 0.85
**Reason:** ANGPTL8 is a well-established regulator of triglyceride metabolism and has been implicated in lipid homeostasis, particularly in association with triglycerides. Its role in lipid regulation through inhibition of lipoprotein lipase makes it a strong candidate for the causal gene in this locus.

GWAS: LDL

Genes in locus: {ZNF45}, {ZNF227}, {ZNF229}, {PVR}, {BCAM}, {RELB}, {EXOC3L2}, {FOSB}, {GIPR}, {ZNF221}, {ZNF233}, {ZNF180}, {BCL3}, {APOC4}, {NKPD1}, {KLC3}, {VASP}, {QPCTL}, {ZNF155}, {ZNF235}, {CEACAM22P}, {CBLC}, {CLASRP}, {MARK4}, {RTN2}, {SNRPD2}, {LOC101928063}, {ZNF285}, {IGSF23}, {NECTIN2}, {GEMIN7}, {CKM}, {PPM1N}, {FBXO46}, {ZNF230}, {ZNF112}, {MIR4531}, {TOMM40}, {TRAPPC6A}, {ERCC1}, {GPR4}, {SIX5}, {ZNF222}, {CEACAM20}, {MIR8085}, {CLPTM1}, {ERCC2}, {OPA3}, {BHMG1}, {ZNF223}, {CEACAM19}, {APOE}, {PPP1R37}, {PPP1R13L}, {EML2}, {SYMPK}, {ZNF284}, {CEACAM16}, {APOC2}, {BLOC1S3}, {CD3EAP}, {MIR330}, {FOXA3}, {ZNF224}, {APOC1}, {MIR6088}, {MIR642A}, {LOC100379224}, {APOC1P1}, {EML2-AS1}

**Likely causal gene:** **APOE**
**Confidence:** 0.9
**Reason:** APOE is well-established in lipid metabolism, particularly influencing LDL cholesterol levels. Numerous GWAS studies have implicated APOE variants in lipid traits, making it a strong candidate for this locus. Although other genes could play a role, APOE's direct link to LDL is well-documented.

GWAS: total cholesterol

Genes in locus: {ZNF45}, {ZNF227}, {ZNF229}, {PVR}, {BCAM}, {RELB}, {EXOC3L2}, {FOSB}, {GIPR}, {ZNF221}, {ZNF233}, {ZNF180}, {BCL3}, {APOC4}, {NKPD1}, {KLC3}, {VASP}, {QPCTL}, {ZNF155}, {ZNF235}, {CEACAM22P}, {CBLC}, {CLASRP}, {MARK4}, {RTN2}, {SNRPD2}, {LOC101928063}, {ZNF285}, {IGSF23}, {NECTIN2}, {GEMIN7}, {CKM}, {PPM1N}, {FBXO46}, {ZNF230}, {ZNF112}, {MIR4531}, {TOMM40}, {TRAPPC6A}, {ERCC1}, {GPR4}, {SIX5}, {ZNF222}, {CEACAM20}, {MIR8085}, {CLPTM1}, {ERCC2}, {OPA3}, {BHMG1}, {ZNF223}, {CEACAM19}, {APOE}, {PPP1R37}, {PPP1R13L}, {EML2}, {SYMPK}, {ZNF284}, {CEACAM16}, {APOC2}, {BLOC1S3}, {CD3EAP}, {MIR330}, {FOXA3}, {ZNF224}, {APOC1}, {MIR6088}, {MIR642A}, {LOC100379224}, {APOC1P1}, {EML2-AS1}

**Likely causal gene**: **APOE**
**Confidence**: 0.9
**Reason**: APOE is a well-established gene associated with lipid metabolism and cholesterol regulation. It has been consistently implicated in GWAS studies of cholesterol levels, with specific alleles affecting lipoprotein metabolism and total cholesterol levels. Other genes in the locus are less directly involved in lipid regulation.

GWAS: BMI

Genes in locus: {STK10}, {SH3PXD2B}, {NEURL1B}, {ERGIC1}, {BNIP1}, {STC2}, {LOC285593}, {CPEB4}, {EFCAB9}, {MIR5003}, {LOC100268168}, {NKX2-5}, {BOD1}, {C5orf47}, {UBTD2}, {LOC101928093}, {CREBRF}, {MIR8056}, {LINC01484}, {LOC100288254}, {DUSP1}, {ATP6V0E1}, {LINC01485}, {RPL26L1}, {SNORA74B}

**Likely Causal Gene**: **CREBRF**
**Confidence**: 0.85
**Reason**: CREBRF has been associated with BMI in multiple studies, including in populations with different genetic backgrounds. The gene encodes a transcription factor involved in metabolic regulation, and genetic variants in CREBRF have been shown to affect fat storage and metabolism, making it a strong candidate for BMI.

GWAS: triglycerides

Genes in locus: {GLTSCR1L}, {SRF}, {ZNF318}, {XPO5}, {VEGFA}, {C6orf223}, {SLC29A1}, {LOC105375075}, {RPL7L1}, {CUL9}, {ABCC10}, {RSPH9}, {LINC01512}, {CAPN11}, {CDC5L}, {SUPT3H}, {C6orf226}, {DNPH1}, {DLK2}, {MRPS18A}, {LOC101929705}, {AARS2}, {PTCRA}, {TTBK1}, {TJAP1}, {MRPL14}, {SPATS1}, {CNPY3}, {PTK7}, {CRIP3}, {LRRC73}, {TMEM63B}, {MIR4642}, {CNPY3-GNMT}, {SLC22A7}, {YIPF3}, {LOC101929726}, {GNMT}, {MIR6780B}, {HSP90AB1}, {PEX6}, {POLR1C}, {SLC35B2}, {PPP2R5D}, {POLH}, {MIR4647}, {MEA1}, {GTPBP2}, {NFKBIE}

**Causal Gene:** **VEGFA**
**Confidence:** 0.85
**Reason:** VEGFA is well-documented in influencing lipid metabolism and vascular health, both of which are directly related to triglyceride levels. It has been implicated in various GWAS studies on lipids, particularly in association with cardiovascular traits, making it a strong candidate for the triglyceride locus.

GWAS: HDL

Genes in locus: {ZNF45}, {ZNF227}, {ZNF229}, {PVR}, {BCAM}, {RELB}, {EXOC3L2}, {FOSB}, {GIPR}, {ZNF221}, {ZNF233}, {ZNF180},

{BCL3}, {APOC4}, {NKPD1}, {KLC3}, {VASP}, {QPCTL}, {ZNF155}, {ZNF235}, {CEACAM22P}, {CBLC}, {CLASRP}, {MARK4}, {RTN2}, {SNRPD2}, {LOC101928063}, {ZNF285}, {IGSF23}, {NECTIN2}, {GEMIN7}, {CKM}, {PPM1N}, {FBXO46}, {ZNF230}, {ZNF112}, {MIR4531}, {TOMM40}, {TRAPPC6A}, {ERCC1}, {GPR4}, {SIX5}, {ZNF222}, {CEACAM20}, {MIR8085}, {CLPTM1}, {ERCC2}, {OPA3}, {BHMG1}, {ZNF223}, {CEACAM19}, {APOE}, {PPP1R37}, {PPP1R13L}, {EML2}, {SYMPK}, {ZNF284}, {CEACAM16}, {APOC2}, {BLOC1S3}, {CD3EAP}, {MIR330}, {FOXA3}, {ZNF224}, {APOC1}, {MIR6088}, {MIR642A}, {LOC100379224}, {APOC1P1}, {EML2-AS1}

**Likely Causal Gene**: **APOE**

**Confidence**: 0.9

**Reason**: APOE is a well-established gene involved in lipid metabolism and HDL regulation. Variants in APOE have been strongly associated with HDL cholesterol levels in multiple studies. Its functional role in lipid transport makes it a prime candidate in this GWAS locus.

GWAS: height

Genes in locus: {ARMC9}, {NMUR1}, {PTMA}, {NPPC}, {MIR562}, {ALPP}, {TIGD1}, {GIGYF2}, {NEU2}, {MIR4777}, {TEX44}, {COPS7B}, {DIS3L2}, {ALPI}, {EFHD1}, {C2orf82}, {INPP5D}, {B3GNT7}, {MIR1244-1}, {ECEL1P2}, {KCNJ13}, {NCL}, {MIR1244-3}, {ALPPL2}, {NGEF}, {SNORA75}, {MIR1244-4}, {ECEL1}, {LOC101928881}, {SNORD20}, {MIR1244-2}, {PRSS56}, {SNORD82}, {PDE6D}, {CHRND}, {LINC00471}, {MIR1471}, {CHRNG}, {MIR5001}, {EIF4E2}

**Likely Causal Gene: PRSS56**

**Confidence: 0.8**

**Reason:** PRSS56 has been implicated in skeletal growth and development in previous studies. Variants in PRSS56 are associated with height, and it plays a role in regulating bone formation, making it a strong candidate for height-related GWAS loci.

GWAS: total cholesterol

Genes in locus: {LOC101929011}, {SIK3}, {TAGLN}, {DSCAML1}, {BUD13}, {PAFAH1B2}, {ZPR1}, {SIDT2}, {APOA5}, {LOC100652768}, {APOA4}, {PCSK7}, {APOC3}, {RNF214}, {APOA1}, {BACE1}, {APOA1-AS}, {BACE1-AS}, {CEP164}

**Likely causal gene**: **APOA5**

**Confidence**: 0.9

**Reason**: APOA5 is well-documented as a key regulator of lipid metabolism, particularly cholesterol levels. Variants in APOA5 are strongly associated with increased triglyceride levels, and it plays a direct role in lipid homeostasis. Its function in cholesterol metabolism and the established GWAS associations make it a likely causal gene.

GWAS: triglycerides

Genes in locus: {ZNF45}, {ZNF227}, {ZNF229}, {PVR}, {BCAM}, {RELB}, {EXOC3L2}, {FOSB}, {GIPR}, {ZNF221}, {ZNF233}, {ZNF180}, {BCL3}, {APOC4}, {NKPD1}, {KLC3}, {VASP}, {QPCTL}, {ZNF155}, {ZNF235}, {CEACAM22P}, {CBLC}, {CLASRP}, {MARK4}, {RTN2}, {SNRPD2}, {LOC101928063}, {ZNF285}, {IGSF23}, {NECTIN2}, {GEMIN7}, {CKM}, {PPM1N}, {FBXO46}, {ZNF230}, {ZNF112}, {MIR4531}, {TOMM40}, {TRAPPC6A}, {ERCC1}, {GPR4}, {SIX5}, {ZNF222}, {CEACAM20}, {MIR8085}, {CLPTM1}, {ERCC2}, {OPA3}, {BHMG1}, {ZNF223}, {CEACAM19}, {APOE}, {PPP1R37}, {PPP1R13L}, {EML2}, {SYMPK}, {ZNF284}, {CEACAM16}, {APOC2}, {BLOC1S3}, {CD3EAP}, {MIR330}, {FOXA3}, {ZNF224}, {APOC1}, {MIR6088}, {MIR642A}, {LOC100379224}, {APOC1P1}, {EML2-AS1}

**Likely Causal Gene:** **APOC4**
**Confidence:** 0.85
**Reason:** APOC4 is part of the apolipoprotein family, which is closely associated with lipid metabolism and triglyceride regulation. Its role in lipid transport and potential impact on triglyceride levels is well-documented in the literature, making it a likely candidate for influencing triglyceride levels in this GWAS locus.

GWAS: triglycerides

Genes in locus: {OTOF}, {CENPA}, {MAPRE3}, {ATRAID}, {NRBP1}, {GPN1}, {RBKS}, {LOC100505736}, {PLB1}, {C2orf70}, {DPYSL5}, {KHK}, {TRIM54}, {FNDC4}, {MRPL33}, {BRE}, {CIB4}, {TMEM214}, {CAD}, {KRTCAP3}, {LINC01460}, {MIR4263}, {LOC100505716}, {KCNK3}, {AGBL5-AS1}, {UCN}, {GCKR}, {BRE-AS1}, {FLJ31356}, {SLC35F6}, {AGBL5}, {SLC30A3}, {C2orf16}, {FOSL2}, {OST4},

{DNAJC5G}, {ZNF512}, {EMILIN1}, {MPV17}, {CCDC121}, {CGREF1}, {GTF3Cf2}, {SUPT7L}, {ABHD1}, {EIF2B4}, {SLC4A1AP}, {PREB}, {SNX17}

**Causal Gene**: **GCKR**
**Confidence**: 0.9
**Reason**: GCKR is strongly associated with triglyceride levels in several GWAS studies. It encodes a protein that regulates glucose metabolism and is involved in hepatic lipid synthesis, linking it directly to triglyceride regulation. Multiple studies support GCKR's involvement in lipid traits, making it the most likely causal gene in this locus.

GWAS: triglycerides

Genes in locus: {LOC101929011}, {SIK3}, {TAGLN}, {DSCAML1}, {BUD13}, {PAFAH1B2}, {ZPR1}, {SIDT2}, {APOA5}, {LOC100652768}, {APOA4}, {PCSK7}, {APOC3}, {RNF214}, {APOA1}, {BACE1}, {APOA1-AS}, {BACE1-AS}, {CEP164}

**Causal Gene**: **APOA5**
**Confidence**: 0.9
**Reason**: APOA5 is a well-established gene linked to triglyceride levels, with multiple studies showing its influence on lipid metabolism. Variants in APOA5 have been consistently associated with altered triglyceride concentrations in both GWAS and functional studies. Its role in lipoprotein metabolism makes it a strong candidate.
